# The CANDID Study: impact of COVID-19 on critical care nurses and organisational outcomes: implications for the delivery of critical care services. A questionnaire study before and during the pandemic

**DOI:** 10.1101/2022.11.16.22282346

**Authors:** Louise McCallum, Janice Rattray, Beth Pollard, Jordan Miller, Alastair Hull, Pam Ramsay, Lisa Salisbury, Teresa Scott, Stephen Cole, Diane Dixon

## Abstract

**Objective:** To use a model of occupational stress to quantify and explain the impact of working in critical care during the Covid-19 pandemic on critical care nurses and organisational outcomes.

**Participants:** Critical care nurses (CCNs) who worked in the UK NHS between January to November 2021 (n=461).

**Methods:** A self-reported survey measured the components of the Job-Demand Reward model of occupational stress. Job-demands, job-resources, health impairment (mental health (GHQ-12), burnout (MBI), PTSD symptoms (PCL-5)), work engagement and six organisational outcomes (commitment, job satisfaction, changing jobs, certainty about the future, quality of care, patient safety) were measured. Data were compared to baseline data (n=557) collected between April to October 2018. Regression analyses identified predictors of health impairment, work engagement and organisational outcomes.

**Findings:** Compared to 2018, CCNs were at elevated risk of probable psychological distress (GHQ-12, OR 6.03 [95% C.I. 4.75 to 7.95]; burnout emotional exhaustion, OR 4.02 [3.07 to 5.26]; burnout depersonalisation, OR 3.18 [1.99 to 5.07]; burnout accomplishment, OR 1.53 [1.18 to 1.97]). A third of CCNs reported probable PTSD. Job demands predicted psychological distress and job demands increased during the pandemic. Resources reduced the negative impact of job demands on psychological distress, but this moderating effect of resources was not observed at higher levels of demand. CCNs were less engaged in their work. Job and personal resources predicted work engagement and were reduced during the pandemic. All six organisational outcomes were impaired. Lack of resources, especially reduced learning opportunities, lack of focus on staff wellbeing, and reduced focus on quality predicted worse organisational outcomes.

**Conclusions:** The NHS needs to prioritise the welfare of CCNs, implement workplace change/planning, and support them to recover from the pandemic. The NHS is struggling to retain CCNs and, unless staff welfare is improved, quality of care and patient safety will likely decline.

## Background

The COVID-19 pandemic inflicted unprecedented and sustained pressure on healthcare systems across the world, the consequences of which were felt most by healthcare workers. Critical care services were in huge demand, indeed media reports during the pandemic described critical care units as the ‘eye of the storm’. These clinical environments were subject to rapid change with the addition of beds and satellite units to cope with surges and super surges in admissions to provide care for the sickest patients. Critical care nursing skills were crucial, this workforce was stretched, and a number of studies have highlighted the consequences including their increased risk of psychological distress and burnout.^1 2^ Yet, the experiences and impact on these nurses and on the healthcare organisations are not fully understood. How an organisation supports its workforce, especially at times of increased demand is key to staff wellbeing, the ability to retain staff and achievement of its aims. We need also to recognise the enduring nature of psychological distress and posttraumatic stress symptoms, which can be difficult to treat, and increase staff sickness and turnover. Taken together, this emphasises the importance of supporting staff both during, and as we emerge from the pandemic for the benefit of individual staff members, quality of critical care services, and ultimately outcomes for patients and their relatives.

Understanding the complex relationships between work environment and individual and organisational outcomes is facilitated in this study by the application of a theoretical model of occupational stress. In 2018 we applied the Job Demand-Resource Model (JD-R)^3^ (Figure 1) to examine the critical care work environment and its impact on Critical Care Nurses (CCNs) and organisational outcomes.^4^ The JD-R model allowed us to measure, understand and test a range of individual factors (personal resources), work environment and job characteristics (job-demands and job-resources) and their association with either negative (e.g. health impairment) or positive (e.g. work engagement) outcomes for CCNs, and importantly organisational outcomes (commitment, job satisfaction, staff turnover, patient safety culture and quality of care). The current study uses this pre-pandemic (2018) data as a unique baseline to report: (1) the impact of the pandemic on CCNs, (2) predictors of CCN work-related stress and work engagement, and (3) the impact on organisational outcomes as a result of the impact of the pandemic on CCNs.

**Fig. 1.**
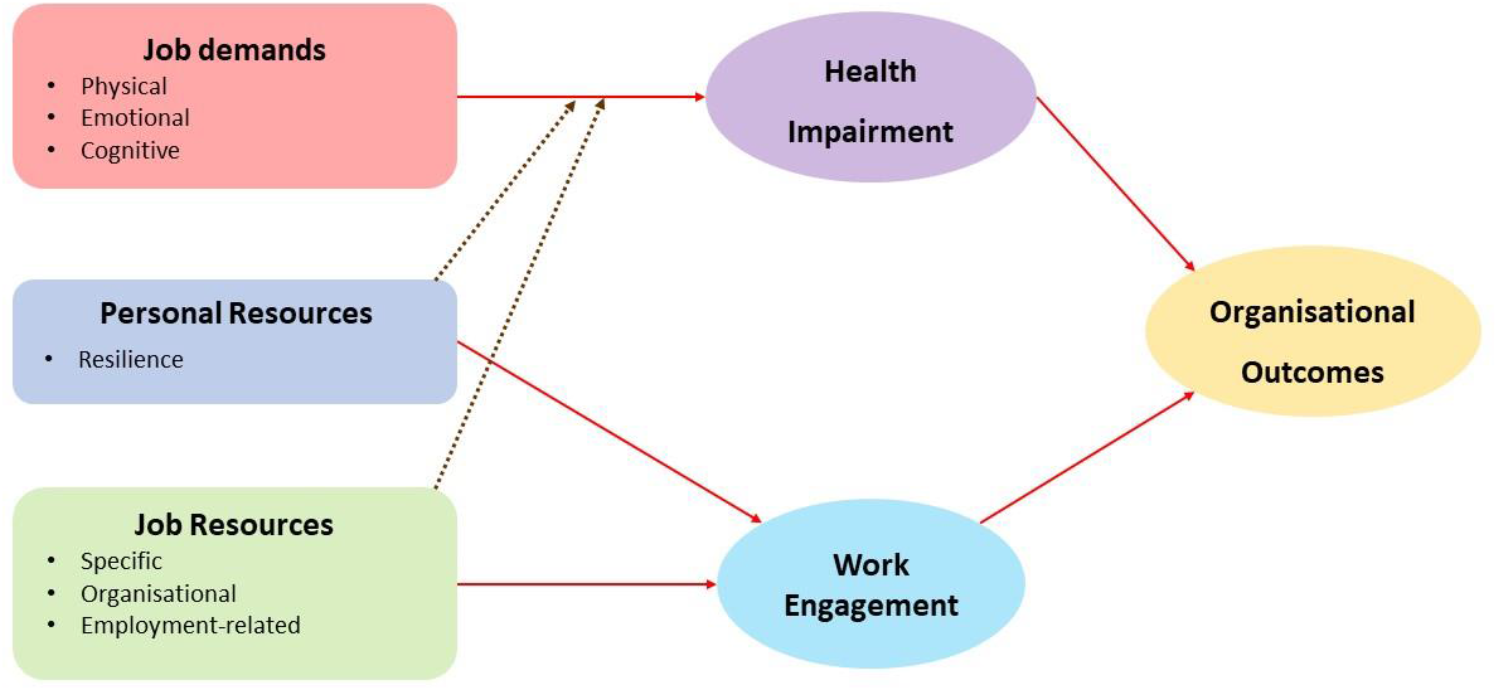
Job Demand-Resource Model

## Methods

Both the current pandemic and pre-pandemic studies involved survey administration. Full details of the CANDID (pandemic) study are in the published study protocol.^5^ The 2018 (pre-pandemic) data were collected between April-October 2018, and the pandemic data between January 2021-March 2022.

### Participants and Recruitment

Participants were CCNs employed within critical care units caring for level three patients across NHS Scotland, two English NHS Trusts and one Welsh NHS Board. Inclusion criteria were Nursing and Midwifery Council registered nurses with substantive contracts in critical care. The 2018 survey recruited from NHS Scotland only. A local study champion in each unit promoted the study and distributed the survey. Participants chose to complete either a paper-based or online survey. Survey completion indicated consent. No individual identifiers were included, although individual critical care units could be.

### Measures

Standard measures with established reliability and validity were used where possible and are listed in Table 1 (full details in Appendix, Table A1). They represent all elements of the JD-R model. The same measures were used in the 2018 survey, except commitment, supporting/communication with relatives and post-traumatic stress symptoms.

**Table 1:**
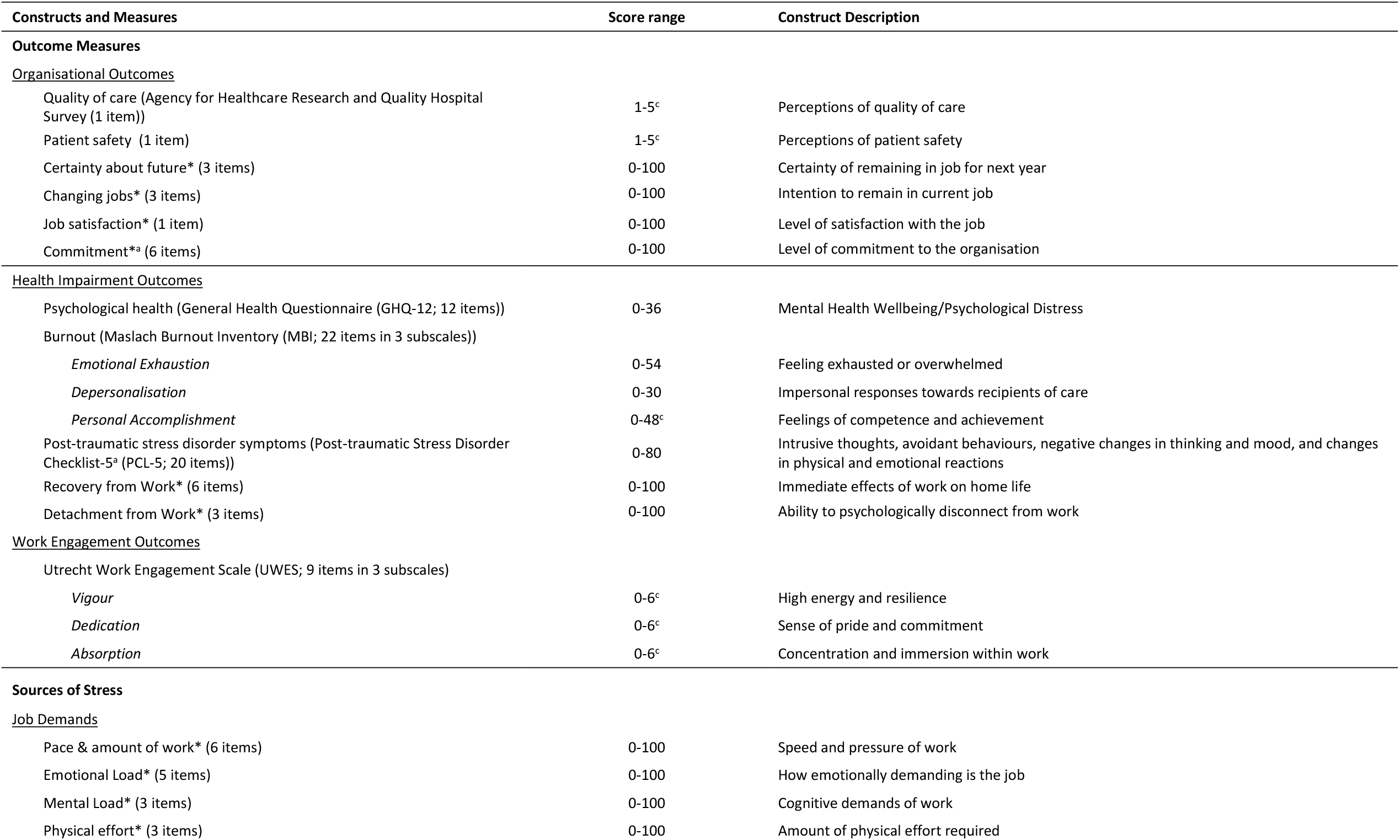
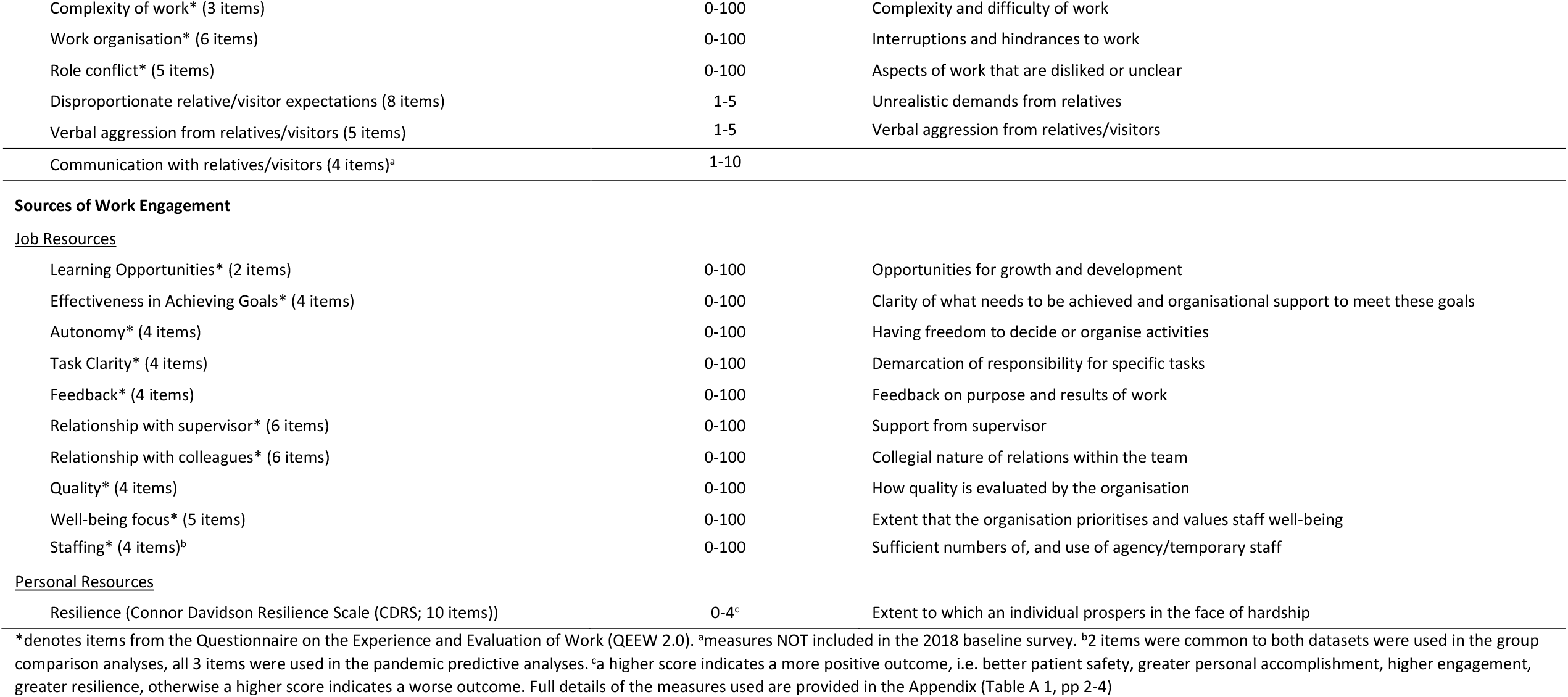
Survey measures and constructs.

### Demographics

A range of demographic, and professional details were collected, including: age, gender, ethnicity, caring responsibilities (children and adults), relationship status, professional details (e.g. years of nursing/CCN experience).

### Sources of Stress (Job-demands) and Work Engagement (Job and Personal Resources)

Nine job-demands and ten job-resources were measured using the QEEW 2.0^6^, and the customer-related social stressors scale^7^ and four items assessing communication with relatives. Personal resources (personal resilience) was measured using the Connor Davidson Resilience Scale (CD-RISC).^8^ Importantly, resilience is a personal attribute that can be viewed as an interaction between a person and their environment i.e., a work environment can either support or undermine a person’s resilience.

### Outcome Measures

The JD-R model specifies three outcomes: i) health impairment, ii) work engagement, and iii) organisational outcomes. Health impairment was operationalised as Burnout Syndrome (Maslach Burnout Inventory (MBI) for Health Services)^9^, Mental Health Well-Being (General Health Questionnaire (GHQ)-12)^10^, Posttraumatic Stress Symptoms (Posttraumatic stress disorder checklist (PCL-5))^11^ and Detachment and Recovery from Work (Questionnaire on the Experience of Work (QEEW2.0)).^6^ The MBI assesses three elements of burnout: ‘emotional exhaustion’, ‘depersonalisation’ and ‘personal accomplishment’. The GHQ-12 is designed to detect current state of psychological distress (social dysfunction, anxiety, depression, and loss of confidence). The PCL-5 assesses intrusive thoughts, avoidant behaviours, negative changes in thinking and mood, changes in physical and emotional reactions, corresponding to DSM-5 symptom criteria for PTSD. Work engagement was measured using the Utrecht Work Engagement Scale, which has three subscales; vigour, dedication and absorption^12^. Six organisational outcomes were assessed, certainty about future, changing jobs, job satisfaction, commitment (QEEW 2.0), quality of care (Agency for Healthcare Research and Quality Hospital Survey)^13^ and, patient safety (Perceptions of Care Undone).^14^

### Data Analysis

Data analysis was performed using IBM SPSSv28. For multi-item measures, treatment of missing values was in accordance with the QEEW methodology as this was the majority of measures, and for consistency this method was applied to all measures. The QEEW method is if an individual has missing values on less than two thirds of items within a scale then the mean of the values that were answered is substituted for the missing values. If more than two thirds of the items were missing, then the scale for that individual was not included in the analysis. If a single item measure was missing, then it was not included in the analysis (See Appendix, Table A2 for details).

Analysis of variance (ANOVA) was used to test differences in outcome means between CCN’s before and during the pandemic. The assumption of equal variances was explored using Levene’s test. Where this indicated lack of equal variances, Welch’s F was reported and Omega squared was used to assess effect size; both were selected for robustness^15^. Logistic regression was used to obtain odds ratios for stress outcomes dichotomised as probable caseness/not according to recommended cut-off scores. A sample size of 500 provides adequate power (80%) to detect a small effect in the GHQ-12 and the estimated prevalence of PTSD (24%) with a precision of 0.035 and confidence of 95%.

Standard linear regression was used to explore outcome predictors. In the first step nursing experience and demographic variables that were significantly correlated with the dependent variable were entered. In the second/third step the relevant JD-R model subscales were entered, i.e. for each Stress outcome the Job-demand subscales were entered; for Organisational outcomes at step 2, the Job-demand and Resource subscales were added and at step 3 the health impairment and work engagement subscales were entered. Multicollinearity was checked using the variance inflation factor (VIF). Normality of residuals was examined by P-P plots and scatterplots were explored to check for homoscedasticity. The significance of the variance contribution of each step was reported together with the R squared change value. Standardised and unstandardised coefficients, t values and significance of the variables within each model were reported.

Nurse experience has been shown to be predictive of outcome^16 17^ and hence was included in all analyses. Nurse experience was also highly correlated with age and critical care experience; to avoid multicollinearity only nurse experience was entered in the analysis. The other demographic variables were gender, pay band, relationship status, childcare, adult care, full/part time, contracted hours.

None of the three standard measures of stress (GHQ-12, MBI and PCL-5) are diagnostic however, both GHQ-12 and PCL-5 have cut-off scores for likelihood of a clinical diagnosis. The GHQ-12 offers two cut-off scores (three and four), we applied both^18^. MBI cut off scores are ≥27 for Emotional Exhaustion, ≥14 for Depersonalisation and between <=30 for Personal Accomplishment; the PCL-5 cut off score is ≥31.^11^

The ability of Job and Personal resources to moderate the relationship between demands and health outcomes was assessed. First, Confirmatory Factor Analysis was carried out using SPSS AMOS to explore Job Demands latent factor to assess whether a composite measure could be computed for use in the moderation analysis. Standard fit indices were explored, acceptable fit was indicated by CFI >0.9, TLI>0.9, RMSEA<0.08 and NFI>0.9. If an acceptable model was found, then imputation was used to create a composite job demand score. Then, Ordinary Least Square regressions were carried to explore moderation. To establish moderation, interaction variables were created by multiplying the independent variable (job demand factor) by each of the potential moderators (i.e. the 11 resource variables). For each health outcome, job demand, the resource and the resource x job demand interaction were entered into a regression (total of 55 regressions). Where a significant interaction was found, this was further probed using simple slopes analysis implemented by the HAYES PROCESS macro in SPSS. A buffering effect would result if as Job Demands increase, the positive effect of the moderator reduces the effect of Job Demands on health outcomes.

### Ethical approval

The study was approved by the Ethical Review Board for the School of Medicine, Medical Sciences and Health at the University of Aberdeen (CERB/2020/10/1993).

### Patient and Public Involvement

Prior to the start of data collection, six CCNs, who were working during the pandemic, were provided with a draft of the study questionnaire. The CCNs were interviewed to sense check questionnaire content and provide feedback in relation to its ease of self-completion and perceived omissions in questionnaire content. The questionnaire was modified accordingly and sent to the study steering group for feedback. The steering group included a retired CCN, the Chair of the British Association of Critical Care Nurses, the Chief Nurse for Research and Development at NHS Lothian, and the Chair of the UK Critical Care Research Group, who also sits on the Intensive Care Society council and the UK Critical Care Nursing Alliance. Two authors and members of the study team (TS and SC) worked in critical care during the pandemic and have provided input throughout. A local unit contact/champion from each participating unit provided study support by promoting and distributing the survey. Preliminary analyses of the study findings were presented to two stakeholder workshops that included CCNs, and senior managers and participants provided feedback on the results. We continue to present the study findings to staff from the Health Boards and Trusts that took part in the study and to receive feedback from them.

### Data Sharing

Requests for access to the data for analyses not contained in the study protocol should be directed to the corresponding authors.

## Results

### Participant characteristics

Table 2 shows the demographic details of the 2018 and pandemic samples. Five hundred and fifty-seven CCNs participated in the 2018 study: a response rate of 47%. The sample size for the pandemic survey was 461. In Scotland the response rate was 32.1%; response rates in England and Wales were substantially lower, but difficult to determine because denominators for these sites could not be identified accurately. All 2018 surveys were paper-based; for the pandemic study, 75% were paper-based and 25% online. The 2018 sample and the pandemic sample did not differ in age, gender, or years of critical care experience.

**Table 2:**
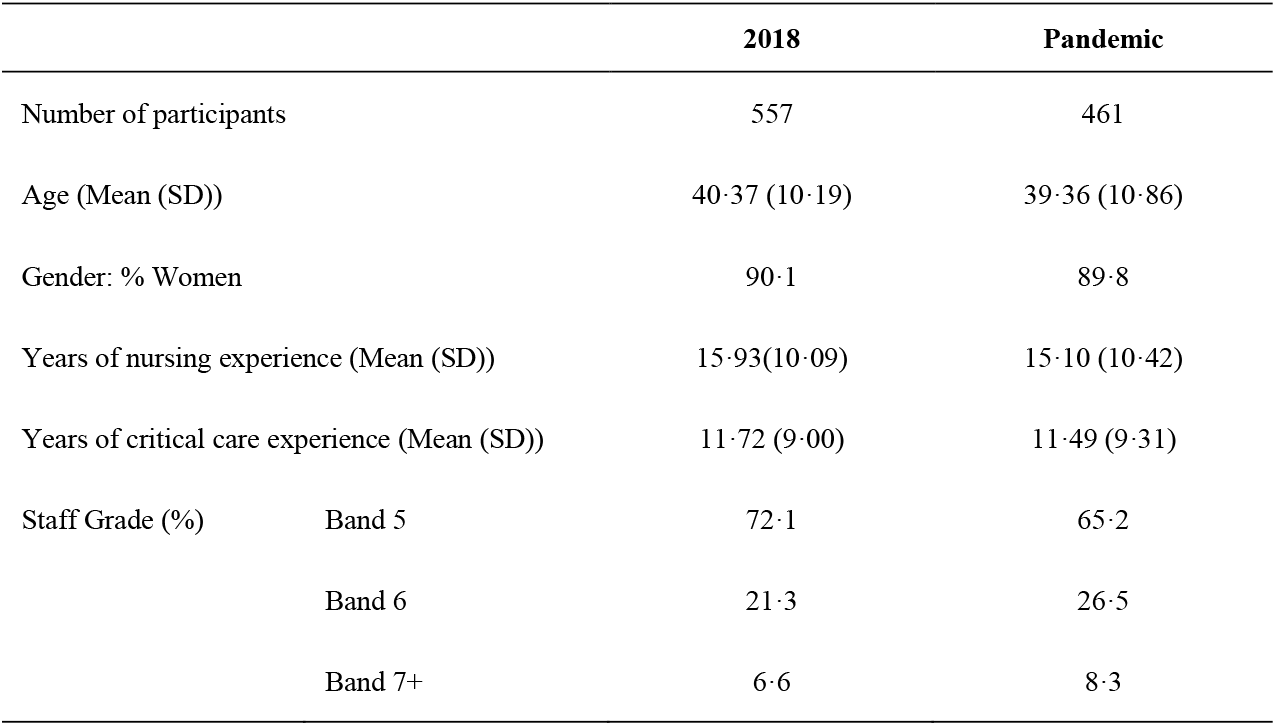
Sample characteristics.

|  |  | 2018 | Pandemic |
| --- | --- | --- | --- |
| Number of participants |  | 557 | 461 |
| Age (Mean (SD)) |  | 40.37 (10.19) | 39.36 (10.86) |
| Gender: % Women |  | 90.1 | 89.8 |
| Years of nursing experience (Mean (SD)) |  | 15.93(10.09) | 15.10 (10.42) |
| Years of critical care experience (Mean (SD)) |  | 11.72 (9.00) | 11.49 (9.31) |
| Staff Grade (%) | Band 5 | 72.1 | 65.2 |
|  | Band 6 | 21.3 | 26.5 |
|  | Band 7+ | 6.6 | 8.3 |

Impact of the pandemic on health impairment, work engagement and organisational outcomes Table 3 reports the levels of each of the three outcomes, (health impairment, work engagement and organisational outcomes) in 2018 and during the pandemic; all were adversely impacted. Health impairment was worse during the pandemic compared to 2018. Scores on symptoms of psychological distress (GHQ-12), burnout (MBI), recovery and detachment after work were significantly higher.

**Table 3:** Levels of Health Impairment, Work Engagement and Organisational Outcomes reported by CCNs in 2018 and during the pandemic.

| Outcome | Mean Score (SD) |  | F (df) | Effect size (Omega2) |
| --- | --- | --- | --- | --- |
|  | 2018 | Pandemic |  |  |
| HEALTH IMPAIRMENT |  |  |  |  |
| Mental Health |  |  |  |  |
| GHQ-12 | 12.52 (5.28) | 18.50 (6.83) | F^(1, 855.69)=236.47*** | 0.20 |
| Burnout |  |  |  |  |
| MBI: Emotional exhaustion | 20.04 (9.81) | 27.85 (11.46) | F^(1, 905.95)=132.60*** | 0.12 |
| MBI: Depersonalisation | 4.97 (4.43) | 6.75 (5.70) | F^(1, 852.08)=29.88*** | 0.03 |
| MBI: Accomplishment | 33.37 (7.32) | 31.45 (7.33) | F(1, 1011)=17.21*** | 0.02 |
| PTSD symptoms |  |  |  |  |
| PCL-5<sup>a</sup> | .. | 24.18 (17.27) | .. | .. |
| Recovery after work | 48.22 (21.63) | 59.32 (22.48) | F(1, 1013)=63.85*** | 0.06 |
| Detachment after work | 41.17 (18.90) | 50.06 (22.30) | F^(1, 905.18)=45.82*** | 0.04 |
| WORK ENGAGEMENT |  |  |  |  |
| Vigour | 3.12 (1.20) | 2.61 (1.24) | F(1, 1011)=43.38*** | 0.04 |
| Dedication | 4.29 (1.06) | 3.94 (1.22) | F^(1, 916.26)=23.23*** | 0.02 |
| Absorption | 3.63 (1.01) | 3.43 (1.21) | F^(1, 893.42)=7.62** | 0.007 |
| ORGANISATIONAL OUTCOMES |  |  |  |  |
| Commitment to the employing organisation<sup>a</sup> | .. | 55.28 (18.09) | .. | .. |
| Satisfaction with working for the employing organisation | 29.80 (19.04) | 49.95 (24.08) | F^(1, 863.89)=211.32*** | 0.18 |
| Certainty about the future (at work) | 18.62 (21.28) | 30.48 (24.66) | F^(1, 926.75)=64.57*** | 0.06 |
| Desire to change jobs | 41.92 (24.74) | 53.50 (27.59) | F^(1, 931.72)=48.58*** | 0.05 |
| Patient safety | 4.21 (.74) | 3.24 (1.00) | F^(1, 780.04)=282.97*** | 0.23 |
| Quality of care | 3.62 (.55) | 2.78 (.96) | F^(1, 651.24)=266.82*** | 0.23 |

GHQ-12: General Health Questionnaire; MBQ: Maslach Burnout Questionnaire (with three subscales); PCL (Post Traumatic Stress Disorder Checklist for *DSM-5*). <sup>a</sup>not measured in 2018 $F^{\wedge}$ =Welch F, \* $p<0.05$ , \*\* $p<0.01$ , \*\*\* $p<0.001$

Table 4 reports the proportion of CCNs reaching cut-off scores for probable caseness on each health impairment measure. The 2018 survey did not include the PCL-5, which precludes the calculation of a change in risk of reaching threshold for caseness. However, 32.54% of the pandemic sample reached the cut-off score, meaning that if a diagnostic interview for PTSD was undertaken, they would very likely have the diagnosis.

**Table 4:** Proportion of CCNs reaching threshold score for health impairment.

|  | 2018 | Pandemic | Odds ratio | OR 95% CI |
| --- | --- | --- | --- | --- |
| **Mental Health** |  |  |  |  |
| GHQ-12 (cut-off score 3) | 34.59 | 76.14 | 6.03*** | 4.75-7.95 |
| GHQ-12 (cut-off score 4) | 28.11 | 69.20 | 5.75*** | 4.83-7.53 |
| **Burnout** |  |  |  |  |
| MBI: Emotional exhaustion | 23.56 | 55.34 | 4.02*** | 3.07-5.26 |
| MBI: Depersonalisation | 4.86 | 13.97 | 3.18*** | 1.99-5.07 |
| MBI: Accomplishment | 33.39 | 43.36 | 1.53** | 1.18-1.97 |
| **PTSD symptoms** |  |  |  |  |
| PCL-5<sup>a</sup> | .. | 32.54 | .. |  |

Data are percentage of nurses reaching threshold. <sup>a</sup>PCL-5 was not included in the 2018 dataset but for context the prevalence of diagnosed PTSD in rescue workers and ambulance workers is 10% and 11% respectively<sup>19 20</sup>. \*\*p<0.01, \*\*\* p<0.001

CCNs were at significantly elevated risk of reaching a cut-off score for caseness on the GHQ-12 and MBI measures. Applying the GHQ-12 cut-off scores of 3 and 4 respectively, in 2018, 34.59% and 28.11% of CCNs reached the cut-off score during the pandemic this increased to 76.14% and 69.20%. CCNs were approximately six times more likely to reach cut-off for caseness using either cut-off score for the GHQ-12 (OR 6.03 95% CI (4.75 to 7.95) for level 3 cut-off and OR 5.75 95% CI (4.83 to 7.53) for level 4 cut-off scores). In 2018, 23.56%, 4.86% and 33.39% met cut-off scores on the emotional exhaustion, depersonalisation and accomplishment sub-scales of the MBI and these increased to 55.34%, 13.97% and 43.36%, respectively during the pandemic. Again, CCNs were at significantly elevated risk of burnout on all three subscales (OR for emotional exhaustion, depersonalisation and accomplishment were 4.02, 3.18, and 1.53 respectively).

All three indicators of work engagement and all five organisational outcomes, measured before and after the pandemic worsened (Table 3). This indicates CCNs were less satisfied with working for their organisation, less certain about their future, more likely to be thinking about changing jobs, and believed quality of care and patient safety had declined (Table 3). The remaining organisational outcome, commitment to working for their organisation, was not measured in 2018.

### Impact on job demands, job and personal resources

The JD-R model theorises that higher job-demands predict greater health impairment, and more resources predict greater work engagement. The pandemic had a variable impact on job-demands (Table 5). Five of the nine job-demands increased; CCNs experienced a higher emotional load, greater task complexity, greater pace and amount of work, greater role conflict and work was less well organised. Mental load was the highest job-demand in 2018 and remained unaffected by the pandemic, as was physical load and verbal aggression from relatives/visitors. In contrast, the job-demand of disproportionate relative/visitor expectations reduced.

**Table 5:** Levels of job demands reported by CCNs before and during the pandemic.

| Type of Job Demand | Job Demand score<br>(Mean (SD)) |  | 2018 v pandemic | Effect size |
| --- | --- | --- | --- | --- |
|  | 2018 | Pandemic |  |  |
| Pace and amount of work | 38.18 (13.62) | 45.23 (16.16) | F<sup>^</sup>(1,900.22)=55.11*** | 0.05 |
| Emotional load | 50.57 (14.30) | 55.93 (15.32) | F(1, 1012)=33.10*** | 0.03 |
| Mental load | 89.40 (13.92) | 89.60 (14.13) | F(1,1101)=0.05 ns | 0.00 |
| Physical effort | 52.42 (21.12) | 52.10 (21.07) | F(1,1012)=0.06 ns | 0.00 |
| Complexity of work | 31.62 (15.99) | 37.45 (17.84) | F(1,1012)=30.18*** | 0.03 |
| Role conflict | 31.67 (12.40) | 33.32 (13.42) | F(1, 1010)=4.08* | 0.003 |
| Work organisation | 47.70 (12.30) | 51.02 (13.90) | F(1, 1013)=16.27*** | 0.02 |
| Verbal aggression from relatives/visitors | 2.46 (.61) | 2.40 (.64) | F(1,1012)=2.06 ns | 0.001 |
| Disproportionate relative/visitor expectations | 2.87 (.60) | 2.77 (.61) | F(1, 1012)=7.22** | 0.006 |

Data are mean (SD). F<sup>^</sup>=Welch F, \*p<0.05, \*\*p<0.01 \*\*\* p<0.001

There was also an impact on job-resources and on CCNs’ personal resilience. Seven of the ten job-resources were negatively impacted by the pandemic (Table 6). CCNs’ reported reduced prioritisation of their wellbeing by their organisation, felt that the emphasis on quality was not as well prioritised, and lacked organisational support to meet goals. Learning and development opportunities were reduced, staffing levels were perceived as insufficient and relationships with colleagues and supervisors were worse. The remaining three job-resources (feedback on performance, task clarity, and autonomy) were not affected. CCN’s personal resilience, was impaired by the pandemic. Thus, job-demands increased during the pandemic and resources to meet those demands were overwhelmingly diminished.

**Table 6:**
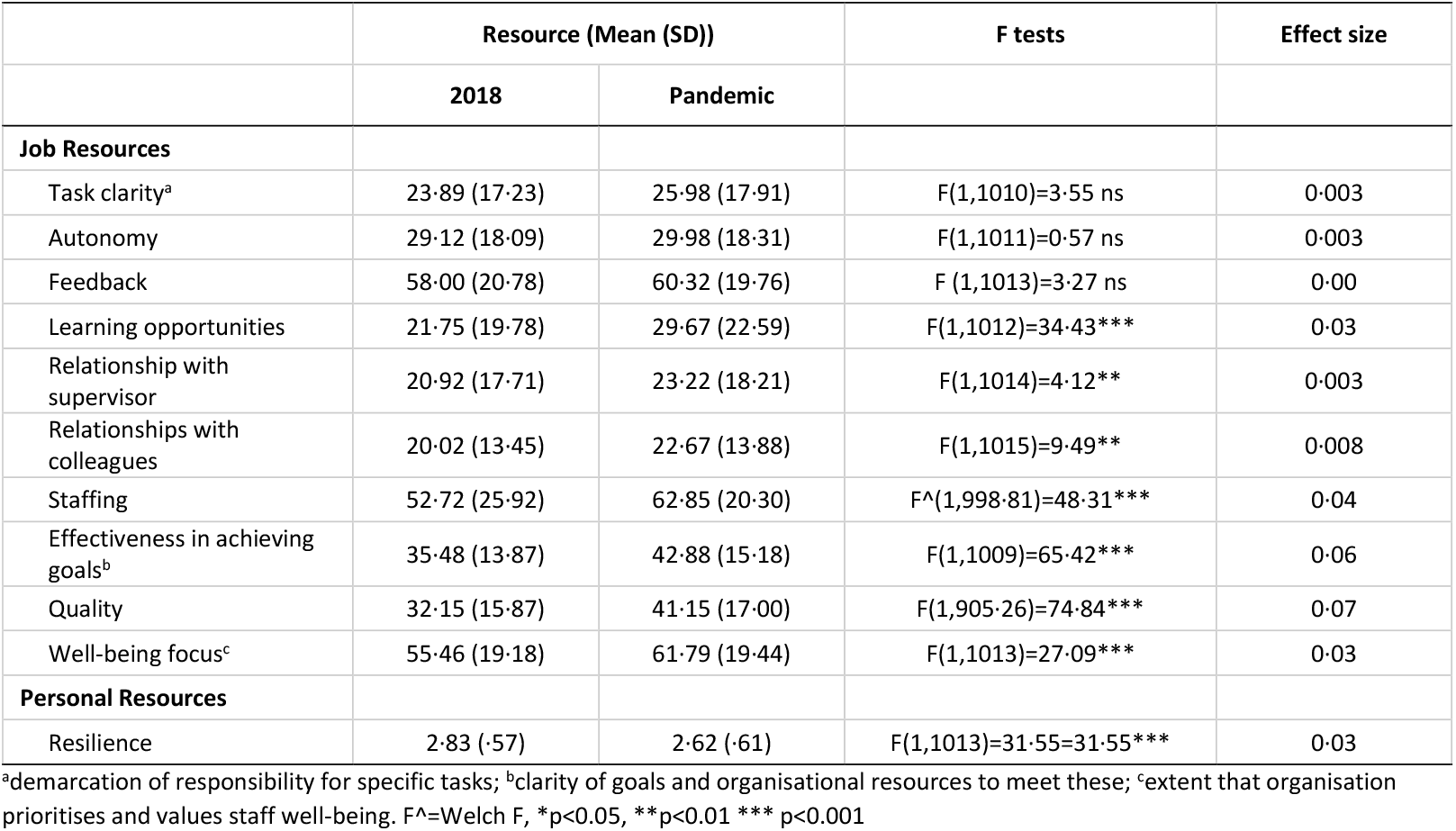
Levels of resources in 2018 and during the pandemic.

### Predictors of health impairment, work engagement and organisational outcomes

The JD-R model identifies job-demands as predictors of stress, and resources as predictors of work engagement. In a series of regression models, we tested the ability of job-demands and resources and demographic factors to predict health impairment and work engagement outcomes. All VIF scores were well below 10, thus multicollinearity did not appear to be a concern. All regression plots indicated that the assumptions of normality and homoscedasticity were acceptable. More nursing experience was protective, being predictive of better outcomes on all health impairment outcomes (Table 7). Emotional load was associated with worse health impairment outcomes on four of the five scales. Higher emotional load was associated with worse psychological distress (GHQ-12), greater emotional exhaustion and depersonalisation (MBI) and higher PTSD symptoms (PCL-5). Poorer work organisation and increased pace and amount of work were associated with worse outcomes on three scales, and challenges with communication with relatives and relatives’ expectations predicted worse outcomes on two of the five scales. Demographic and job-demand variables accounted for over a quarter of the variance in GHQ-12 and depersonalisation, over a third of the variance in PCL-5 scores and almost half of the variance in emotional exhaustion, whereas they only explained an eighth of the variance in personal achievement.

**Table 7:**
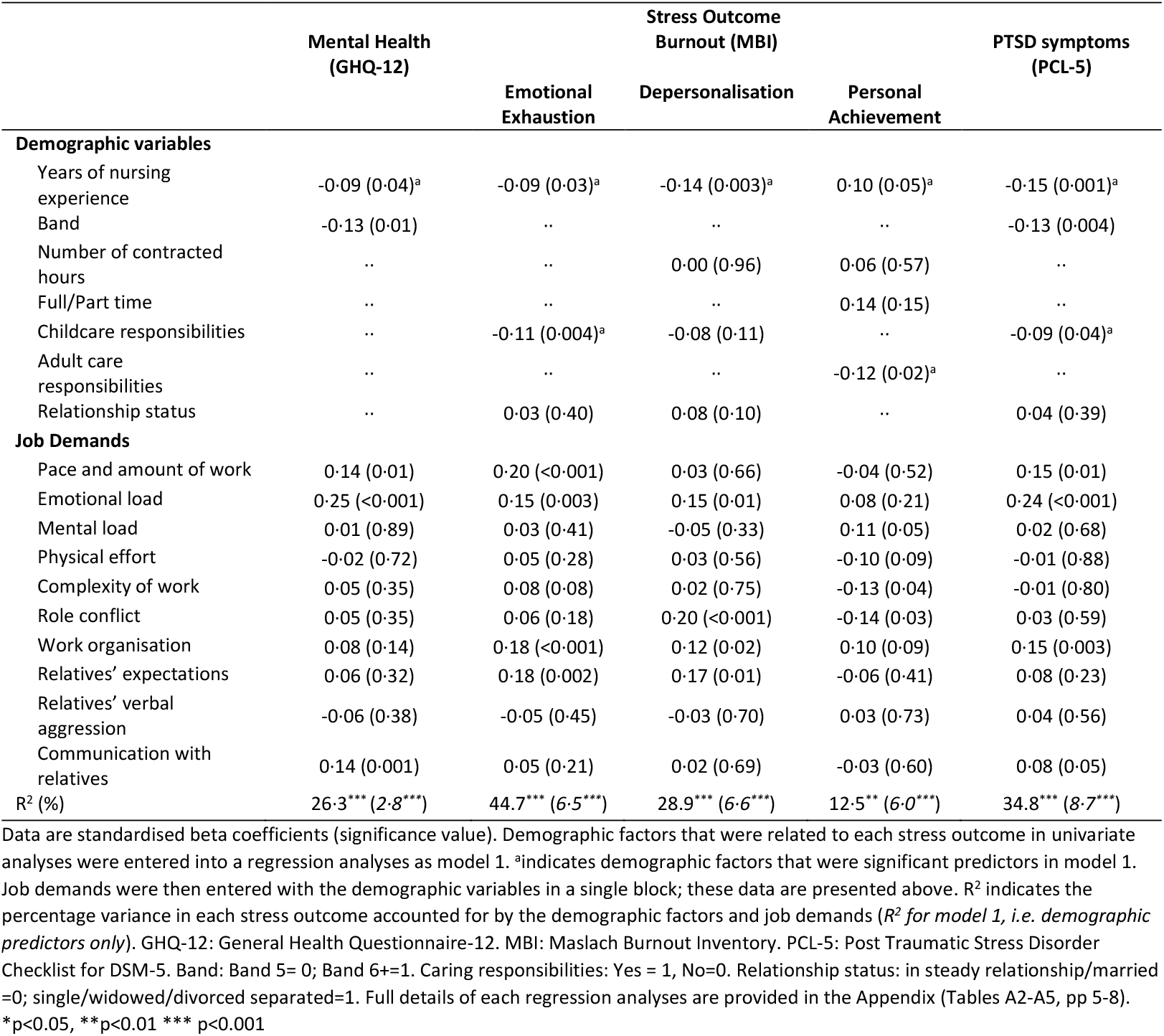
Multivariate model to identify the job demands that predict stress.

|  | Mental Health<br>(GHQ-12) | Stress Outcome<br>Burnout (MBI) |  |  | PTSD symptoms<br>(PCL-5) |
| --- | --- | --- | --- | --- | --- |
|  |  | Emotional<br>Exhaustion | Depersonalisation | Personal<br>Achievement |  |
| Demographic variables |  |  |  |  |  |
| Years of nursing experience | -0.09 (0.04)<sup>a</sup> | -0.09 (0.03)<sup>a</sup> | -0.14 (0.003)<sup>a</sup> | 0.10 (0.05)<sup>a</sup> | -0.15 (0.001)<sup>a</sup> |
| Band | -0.13 (0.01) | .. | .. | .. | -0.13 (0.004) |
| Number of contracted hours | .. | .. | 0.00 (0.96) | 0.06 (0.57) | .. |
| Full/Part time | .. | .. | .. | 0.14 (0.15) | .. |
| Childcare responsibilities | .. | -0.11 (0.004)<sup>a</sup> | -0.08 (0.11) | .. | -0.09 (0.04)<sup>a</sup> |
| Adult care responsibilities | .. | .. | .. | -0.12 (0.02)<sup>a</sup> | .. |
| Relationship status | .. | 0.03 (0.40) | 0.08 (0.10) | .. | 0.04 (0.39) |
| Job Demands |  |  |  |  |  |
| Pace and amount of work | 0.14 (0.01) | 0.20 (<0.001) | 0.03 (0.66) | -0.04 (0.52) | 0.15 (0.01) |
| Emotional load | 0.25 (<0.001) | 0.15 (0.003) | 0.15 (0.01) | 0.08 (0.21) | 0.24 (<0.001) |
| Mental load | 0.01 (0.89) | 0.03 (0.41) | -0.05 (0.33) | 0.11 (0.05) | 0.02 (0.68) |
| Physical effort | -0.02 (0.72) | 0.05 (0.28) | 0.03 (0.56) | -0.10 (0.09) | -0.01 (0.88) |
| Complexity of work | 0.05 (0.35) | 0.08 (0.08) | 0.02 (0.75) | -0.13 (0.04) | -0.01 (0.80) |
| Role conflict | 0.05 (0.35) | 0.06 (0.18) | 0.20 (<0.001) | -0.14 (0.03) | 0.03 (0.59) |
| Work organisation | 0.08 (0.14) | 0.18 (<0.001) | 0.12 (0.02) | 0.10 (0.09) | 0.15 (0.003) |
| Relatives' expectations | 0.06 (0.32) | 0.18 (0.002) | 0.17 (0.01) | -0.06 (0.41) | 0.08 (0.23) |
| Relatives' verbal aggression | -0.06 (0.38) | -0.05 (0.45) | -0.03 (0.70) | 0.03 (0.73) | 0.04 (0.56) |
| Communication with relatives | 0.14 (0.001) | 0.05 (0.21) | 0.02 (0.69) | -0.03 (0.60) | 0.08 (0.05) |
| R<sup>2</sup> (%) | 26.3*** (2.8***) | 44.7*** (6.5***) | 28.9*** (6.6***) | 12.5** (6.0***) | 34.8*** (8.7***) |

Data are standardised beta coefficients (significance value). Demographic factors that were related to each stress outcome in univariate analyses were entered into a regression analyses as model 1. <sup>a</sup>indicates demographic factors that were significant predictors in model 1. Job demands were then entered with the demographic variables in a single block; these data are presented above. R<sup>2</sup> indicates the percentage variance in each stress outcome accounted for by the demographic factors and job demands (*R<sup>2</sup> for model 1, i.e. demographic predictors only*). GHQ-12: General Health Questionnaire-12. MBI: Maslach Burnout Inventory. PCL-5: Post Traumatic Stress Disorder Checklist for DSM-5. Band: Band 5= 0; Band 6+=1. Caring responsibilities: Yes = 1, No=0. Relationship status: in steady relationship/married =0; single/widowed/divorced separated=1. Full details of each regression analyses are provided in the Appendix (Tables A2-A5, pp 5-8). \*p<0.05, \*\*p<0.01 \*\*\* p<0.001

Three of the 11 resources (job and personal) measured predicted work engagement, namely, quality, learning opportunities and resilience; more resources predicted better engagement. None of the demographic factors were predictive (Table 8). These three resources predicted all three measures of work engagement, with resilience being the strongest predictor. Resources accounted for approximately a third of the variability in vigour and dedication and approximately a quarter of the variability in absorption.

**Table 8:** Multivariate model to identify the resources that predict work engagement.

|  | Indicator of work engagement |  |  |
| --- | --- | --- | --- |
|  | Vigour | Dedication | Absorption |
| **Demographic variables** |  |  |  |
| Years of nursing experience | 0.01 (0.85) | 0.04 (0.45) | 0.08 (0.11) |
| Band | .. | .. | -0.01 (0.86) |
| Number of contracted hours | .. | 0.08 (0.11) | 0.11 (0.22) |
| Full/Part time | .. | .. | -0.02 (0.82) |
| Childcare responsibilities | 0.05 (0.20) | 0.06 (0.19) | .. |
| **Job Resources** |  |  |  |
| Task Clarity | -0.03 (0.55) | -0.06 (0.24) | 0.00 (0.94) |
| Autonomy | -0.03 (0.61) | 0.05 (0.34) | -0.02 (0.67) |
| Feedback | -0.07 (0.17) | -0.02 (0.70) | -0.07 (0.16) |
| Relationship with supervisor | -0.05 (0.30) | -0.07 (0.22) | 0.00 (1.00) |
| Relationship with colleagues | -0.03 (0.60) | -0.03 (0.57) | 0.05 (0.38) |
| Staffing | -0.06 (0.24) | -0.06 (0.20) | 0.00 (0.94) |
| Wellbeing | -0.09 (0.06) | -0.04 (0.37) | 0.00 (0.97) |
| Effectiveness achieving goals | 0.08 (0.14) | -0.01 (0.87) | 0.07 (0.23) |
| Quality | -0.15 (0.01) | -0.11 (0.05) | -0.24 (<0.001) |
| Learning Opportunities | -0.12 (0.01) | -0.24 (<0.001) | -0.13 (0.01) |
| **Personal Resources** |  |  |  |
| Resilience | 0.38 (<0.001) | 0.33 (<0.001) | 0.37 (<0.001) |
| R<sup>2</sup> (%) | 33.5*** (2.2**) | 35.1*** (3.5**) | 28.2*** (3.8**) |

Data are standardised beta coefficients (significance value). Demographic factors that were related to each work engagement outcome in univariate analyses were entered into a regression analyses as model 1. <sup>a</sup>indicates demographic factors that were significant predictors in model 1. Resources were then entered with the demographic variables in a single block; these data are presented above. R<sup>2</sup> indicates the percentage variance in each work engagement outcome accounted for by the demographic factors and resources (R<sup>2</sup> for model 1, i.e. demographic predictors only). Full/Part time: Part time =0, Full time=1. Caring responsibilities: Yes = 1, No=0. Full details of each regression analyses are provided in the Appendix (Tables A6-A8, pp 9-11). \*p<0.05, \*\*p<0.01 \*\*\* p<0.001

The JD-R model identifies health impairment and work engagement as predictors of organisational outcomes. However, using latent constructs as predictors does not identify specific actionable predictors. Actionable understanding is required to inform subsequent interventions to support staff. Therefore, a series of regression analyses were performed that allowed job-demands and resources, and work engagement and health impairment measures, to predict organisational outcomes. The results of the regression analyses are shown in Table 9 in the form of a heat map.

**Table 9:**
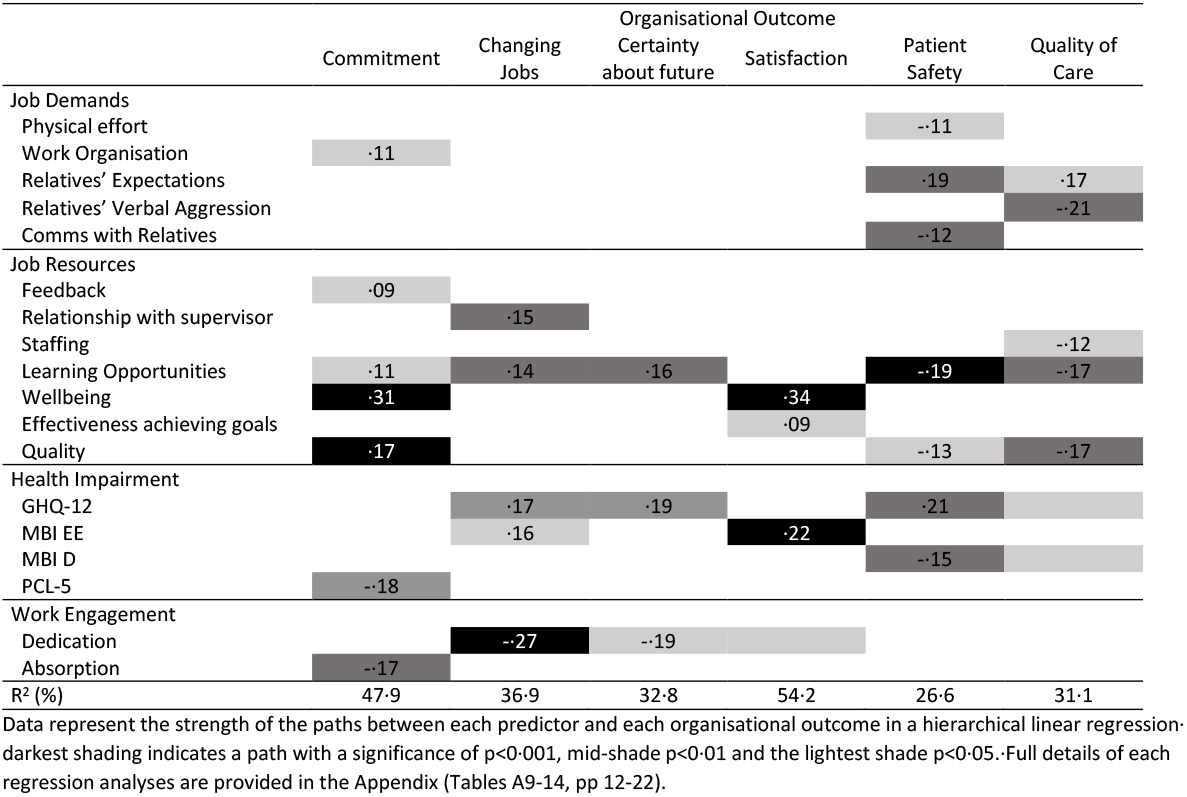
Predictors of organisational outcomes.

All six organisational outcomes were predicted by health impairment, work engagement, individual job-demands and resources. Five of the nine job-demands predicted at least one organisational outcome, whilst seven of the 11 resources were predictive. All three health impairment measures predicted at least one organisational outcome and two of the three indicators of work engagement (dedication and absorption) were predictive. Table 9 indicates that job-resources and health impairments are the more important predictors of organisational outcomes, predicting a greater number of outcomes and being stronger predictors than job-demands or work engagement.

Significant and substantial proportions of the variability in all six organisational outcomes were explained by components of the JD-R model, e.g., approximately half of the variability in job satisfaction and job commitment was accounted for. Some outcomes, e.g., quality of care, patient safety and commitment, were explained by a wide range of predictors, whilst others, e.g., certainty about the future and satisfaction had only a few. Learning opportunities are a resource of note as they strongly predicted five of the six outcomes; they did not predict job satisfaction.

### Moderation of the impact of demands on health impairment by resources

The CFA for job demand indicated a good initial model fit though any subscale with path coefficients less than 0.3 were removed. This resulted in communication with relatives being removed. The resultant model had very good model fit (CFI=0.96; NFI=0.94; TLI=0.94; RMSEA=0.07). Hence, imputation was used to compute the composite job demand score.

Moderators were identified for mental health (GHQ-12), burnout emotional exhaustion (MBI-EE) and PTSD symptoms (PCL-5) (see supplementary file for full details). One job resource, relationship with colleagues, moderated the relationship between demands and mental health (beta=0.17, p≤0.05). Four resources, namely, learning opportunities (beta=-0.17, p≤0.05), prioritisation of well-being (beta=-0.30, p≤0.05), relationships with colleagues (beta=-0.17, p≤0.05) and relationships with supervisor (beta=-0.21, p≤0.001) moderated the relationship between demands and burnout emotional exhaustion. Simple slopes analyses indicated that resources reduced the negative effect of demands on mental health and burnout emotional exhaustion. However, this moderation effect was stronger at lower levels of demand; as demands increased, the moderation effect reduced (see Appendix, Figures A1 to A6). Learning opportunities also potentially moderated the relationship between demands and PTSD symptoms (beta=0.015,). At low levels of demand, learning opportunities mitigated the negative impact of demands on PTSD symptoms, whereas this moderation effect was not apparent at high levels of demand.

## Discussion

This study revealed the considerable adverse impact of the pandemic on the wellbeing of CCNs, particularly in relation to risk for work-related stress and the impact on NHS organisations. Up to three quarters of CCNs are at risk of significant psychological distress, up to half at risk of burnout and a third reported PTSD symptoms at a level that would require formal clinical assessment. Crucially, having pre-COVID-19 comparative data strengthens the importance of these results. These levels of probable mental health impairment represent a significantly elevated risk compared to the 2018 data, and are consistent with rates reported in critical care healthcare workers in the UK^1 21^ and internationally^22^ during the pandemic. CCN risk is three times higher than comparator occupations that are likely to experience traumatic events, namely, rescue workers and ambulance workers^19 20^, with risk factors relating to the trauma, the person, and the environment.

PTSD symptoms are a consequence of traumatic experience, and the trajectory for such psychological responses can be enduring^8^, with significant additional functional impairment.^5^ Burnout is also enduring^23^ and can be contagious through horizontal transfer, therefore prevalence may increase^24^, particularly in new staff. Burnout also has a negative impact on quality of care and career engagement.^25^ Collectively, these findings have implications for a workforce that is still experiencing high levels of clinical activity and attrition within a challenging work environment. It is also likely that the impacts on CCNs reported here will also be relevant to other critical care staff.

A unique aspect of this study was the pre-pandemic data. Through this approach, we were able to unequivocally demonstrate that all indicators of work engagement and organisational outcomes, for which 2018 data were available, were negatively impacted. The worsening of organisational outcomes is of importance for staff retention and may have significant implications for individual hospitals and the NHS nationally. CCNs reported less job satisfaction, were less certain about their future and were more likely to be considering changing jobs, a major concern in relation to current staffing challenges. Staff attrition risks understaffing and potentially negative impact on staff wellbeing and development. In particular, the loss of experienced staff will be detrimental to the support, supervision and retention of staff new to critical care, who in the post pandemic era are increasingly required to fill gaps in staffing. Staff perceptions of reduced quality of care and patient safety are concerning and have implications for patients, their families and staff, and is a potential cause of stress.

CCNs experienced significantly increased job-demands with concurrent reduction in resources. The increase in specific job-demands was perhaps unsurprising. The increased number of patients, greater patient acuity and mortality rates likely contributed to this perception, as was working in satellite units with redeployed staff and delivering care in full PPE. Only one demand decreased, namely, interacting with relatives: that relatives were not allowed into ICU during the pandemic likely explains this finding.

Seven of the 10 job-resources were significantly reduced, alongside the personal resource of resilience. It may be that by assessing personal resilience during the pandemic, individuals had insufficient recovery time. However, resilience depletion may also be an indication of the impact of the ongoing pandemic and continuing high levels of (largely non-COVID) clinical demand; this must be addressed. Emphasis on quality of care and the organisation’s focus on staff wellbeing (organisational job-resources) were worse during the pandemic. This has important ramifications as both resources, together with learning opportunities, were predictive of staff’s work engagement.

The development of effective staff support measures requires the identification and understanding of predictors of health impairment, work engagement and organisational outcomes for staff now and for future pandemic planning. Nursing experience and having caring responsibilities for children were both associated with better mental health. This is clearly complex, given staff’s concern of transmitting COVID to family members, but replicates pre-pandemic findings^26^. It has been suggested that workplace flexibility may explain the relationship between caring responsibilities and workplace stress. Staff with no caring responsibilities outside work, may feel additional pressure to volunteer to cover shifts and work longer hours, whilst staff with caring responsibilities may have a greater degree of flexibility built into their working pattern. ^27^ CCNs experienced a constellation of increased job-demands, each predictive of psychological distress on one or more measure. Social support has been shown to protect against the development of PTSD after exposure to a traumatic event^28^ and this was likely reduced during the pandemic via restrictions on social contact. Relationships worsened between CCNs and their supervisors and colleagues, thereby impairing important sources of social support at work.^29^ Importantly, the ability of resources to mitigate the negative effects of demands on health impairment was reduced at high levels of demand. This suggests that when demands are at their highest, the mental health of CCNs cannot be protected through provision of extra resources, rather, the level of demand must be reduced.

While the loss of supportive resources explains the reduction in work engagement, it also clarifies the impact of the pandemic on organisational outcomes with job-resources and health impairment the strongest and most consistent predictors. The maintenance and promotion of learning opportunities both during and after the pandemic is vital as it was an important resource for CCNs. The key influence of job-resources and how quality and focus on wellbeing were perceived by CCNs will concern senior management teams, but crucially also provides a focus for improving staff wellbeing. Supporting current CCNs is essential to prevent a worsening exodus of highly skilled, experienced and committed staff.

A key strength of this study is the application of a theoretical model allowing the testing of relationships across multiple independent and dependent variables. Participants were recruited across the United Kingdom, enhancing the generalisability of our findings. COVID-19 was a global pandemic, and evidence indicates a role of workplace factors in its impact on healthcare workers globally^30^, suggesting these UK findings have international significance. The data were collected during the height of the pandemic and while the response rates, particularly those from the non-Scotland sites, were less than the baseline study there was group equivalence across the key clinical and demographic variables.

In conclusion, health impairment and work engagement in CCNs was significantly worse than pre-pandemic, and this must be an ongoing issue for healthcare organisations nationally and internationally. As predicted, both of these issues along with organisational outcomes, were associated with increased job-demands but also – and more strongly - with a lack of resources. The importance of job resources perhaps reflect a wider issue of culture, and represents how staff perceive their value, treatment and support. This is key in staff retention and overall quality of care. The implications for CCN recruitment and retention is stark. Moreover, these consequences are ongoing, may worsen and need to be addressed urgently. The influence of job-resources especially in relation to learning opportunities, quality and wellbeing - and their importance to CCNs – must not be underestimated by healthcare organisations. CCNs made a significant contribution to the UK response to the pandemic and their wellbeing now and in the future must be prioritised.

## Data Availability

All data produced in the present study are available upon reasonable request to the authors

## Contributor statement

The corresponding author attests that all listed authors meet authorship criteria and that no others meeting the criteria have been omitted.

## Copyright

The Corresponding Author has the right to grant on behalf of all authors and does grant on behalf of all authors, an exclusive licence on a worldwide basis to the BMJ Publishing Group Ltd to permit this article (if accepted) to be published in BMJ editions and any other BMJPGL products and sublicences such use and exploit all subsidiary rights, as set out in our licence

## Transparency statement

The corresponding author affirms that this manuscript is an honest, accurate, and transparent account of the study being reported; that no important aspects of the study have been omitted; and that any discrepancies from the study as planned (and, if relevant, registered) have been explained.

## Acknowledgements

The authors would like to extend their thanks to all the unit champions who worked to promote the study locally in each unit. We are also very grateful to all the nurses who gave of their time to take part in the development of the study questionnaire and in CANDID itself. The study also benefitted from a very thoughtful Steering Group, whose input improved the study questionnaire and helped develop our dissemination strategy; we are very grateful to them. Finally, we would like to thank the company NURSEM who generously supplied CANDID with hand cream, which was given to each unit as a ‘thank you’ from the CANDID team.

