## Supplementary material for "The CANDID Study: impact of COVID-19 on critical care nurses and organisational outcomes: implications for the delivery of critical care services. A questionnaire study before and during the pandemic": online appendix

**Table A1: Study Measures**

| | Score range | Construct Description | Measure reference or item wording |
|---|---|---|---|
| **Outcome Measures** | | | |
| **Organisational Outcomes** | | | |
| Quality of care (Agency for Healthcare Research and Quality Hospital Survey (1 item)) | 1-5[c] | Perceptions of quality of care | Rockville W, Sorra J, Gray L, et al. AHRQ Hospital Survey on Patient Safety Culture: User's Guide. Rockville, MD: Agency for Healthcare Research and Quality U.S. Department of Health and Human Services 2018. |
| Patient safety (1 item) | 1-5[c] | Perceptions of patient safety | Nurses rated the following statement: *Thinking about the first phase of the COVID-19 pandemic, please give the unit that you work on, an overall grade on patient safety;* on the response scale: Failing, Poor, Acceptable, Very Good, Excellent |
| Certainty about future* (3 items) | 0-100 | Certainty of remaining in job for next year | Veldhoven M, Prins J, van der Laken P, Dijkstra L. QEEW2.0: 42 short scales for survey research on work, well-being and performance; 2015. |
| Changing jobs* (3 items) | 0-100 | Intention to remain in current job | |
| Job satisfaction* (1 item) | 0-100 | Level of satisfaction with the job | |
| Commitment*[a] (6 items) | 0-100 | Level of commitment to the organisation | |
| **Health Impairment Outcomes** | | | |
| Psychological health (General Health Questionnaire (GHQ-12; 12 items)) | 0-36 | Mental Health Wellbeing/Psychological Distress | Goldberg DP, Hillier VF. A scaled version of the General Health Questionnaire. Psychol Med 1979; 9(1): 139-45. |
| Burnout (Maslach Burnout Inventory (MBI; 22 items in 3 subscales)) | | | |
| *Emotional Exhaustion* | 0-54 | Feeling exhausted or overwhelmed | Maslach C, Jackson SE. The measurement of experienced burnout. Journal of Organizational Behavior 1981; 2(2): 99-113. |
| *Depersonalisation* | 0-30 | Impersonal responses towards recipients of care | |
| *Personal Accomplishment* | 0-48[c] | Feelings of competence and achievement | |
| Post-traumatic stress disorder symptoms (Post-traumatic Stress Disorder Checklist-5[a] (PCL-5; 20 items)) | 0-80 | Intrusive thoughts, avoidant behaviours, negative changes in thinking and mood, and changes in physical and emotional reactions | Weathers FW, Litz BT, Keane TM, Palmieri PA, Marx BP, Schnurr PP. The PTSD Checklist for DSM-5 (PCL-5). 2013. www.ptsd.va.gov. |
| Recovery from Work* (6 items) | 0-100 | Immediate effects of work on home life | Veldhoven M, Prins J, van der Laken P, Dijkstra L. QEEW2.0: 42 short scales for survey research on work, well-being and performance; 2015. |
| Detachment from Work* (3 items) | 0-100 | Ability to psychologically disconnect from work | |
| **Work Engagement Outcomes** | | | |

| | | | |
|---|---|---|---|
| Utrecht Work Engagement Scale (UWES; 9 items in 3 subscales) | | | |
| *Vigour* | 0-6[c] | High energy and resilience | Schaufeli WB, Bakker AB, Salanova M. The Measurement of Work Engagement With a Short Questionnaire:A Cross-National Study. Educational and Psychological Measurement 2006; 66(4): 701-16. |
| *Dedication* | 0-6[c] | Sense of pride and commitment | |
| *Absorption* | 0-6[c] | Concentration and immersion within work | |
| **Sources of Stress** | | | |
| **Job Demands** | | | |
| Pace & amount of work* (6 items) | 0-100 | Speed and pressure of work | Veldhoven M, Prins J, van der Laken P, Dijkstra L. QEEW2.0: 42 short scales for survey research on work, well-being and performance; 2015. |
| Emotional Load* (5 items) | 0-100 | How emotionally demanding is the job | |
| Mental Load* (3 items) | 0-100 | Cognitive demands of work | |
| Physical effort* (3 items) | 0-100 | Amount of physical effort required | |
| Complexity of work* (3 items) | 0-100 | Complexity and difficulty of work | |
| Work organisation* (6 items) | 0-100 | Interruptions and hindrances to work | |
| Role conflict* (5 items) | 0-100 | Aspects of work that are disliked or unclear | |
| Disproportionate relative/visitor expectations (8 items) | 1-5 | Unrealistic demands from relatives | Dormann C, Zapf D. Customer-related social stressors and burnout. J Occup Health Psychol. 2004 Jan;9(1):61-82. doi: 10.1037/1076-8998.9.1.61. |
| Verbal aggression from relatives/visitors (5 items) | 1-5 | Verbal aggression from relatives/visitors | |
| Communication with relatives/visitors (4 items)[a] | 1-10 | Communication challenges during the pandemic | Nurses rated how challenging they found the following four statements: 1. *Developing a good relationship with relatives*; 2. *Speaking with relatives remotely*; 3. *Caring for patients in the absence of their relatives*; 4. *Supporting relatives during end of life care*; on a response scale of 1 = not at all challenging through to 10 = extremely challenging. |
| **Sources of Work Engagement** | | | |
| **Job Resources** | | | |
| Learning Opportunities* (2 items) | 0-100 | Opportunities for growth and development | Veldhoven M, Prins J, van der Laken P, Dijkstra L. QEEW2.0: 42 short scales for survey research on work, well-being and performance; 2015. |
| Effectiveness in Achieving Goals* (4 items) | 0-100 | Clarity of what needs to be achieved and organisational support to meet these goals | |
| Autonomy* (4 items) | 0-100 | Having freedom to decide or organise activities | |
| Task Clarity* (4 items) | 0-100 | Demarcation of responsibility for specific tasks | |
| Feedback* (4 items) | 0-100 | Feedback on purpose and results of work | |
| Relationship with supervisor* (6 items) | 0-100 | Support from supervisor | |
| Relationship with colleagues* (6 items) | 0-100 | Collegial nature of relations within the team | |

| | | | |
|---|---|---|---|
| Quality* (4 items) | 0-100 | How quality is evaluated by the organisation | |
| Well-being focus* (5 items) | 0-100 | Extent that the organisation prioritises and values staff well-being | |
| Staffing* (4 items)[b] | 0-100 | Sufficient numbers of, and use of agency/temporary staff | |
| **Personal Resources** | | | |
| Resilience (Connor Davidson Resilience Scale (CDRS; 10 items)) | 0-4[c] | Extent to which an individual prospers in the face of hardship | Connor KM, Davidson JR. Development of a new resilience scale: the Connor-Davidson Resilience Scale (CD-RISC). Depress Anxiety 2003; 18(2): 76-82. |

*denotes items from the Questionnaire on the Experience and Evaluation of Work (QEEW 2.0). [a]measures NOT included in the 2018 baseline survey. [b]2 items were common to both datasets were used in the group comparison analyses, all 3 items were used in the pandemic predictive analyses. [c]a higher score indicates a more positive outcome, i.e. better patient safety, greater personal accomplishment, higher engagement, greater resilience, otherwise a higher score indicates a worse outcome.

#### MISSING DATA

For multi-item measures, treatment of missing values was in accordance with the QEEW methodology as this was the majority of measures and for consistency this method was applied to all measures. The QEEW method is if an individual has missing values on less than two thirds of the items within a scale then the mean of the answered items was substituted for the missing values. If more than two thirds of the items were missing, then it was not included in the analysis. If a single item measure was missing, then it was not included in the analysis.

**Table A2: Missing Data For Each Outcome**

| **Variable** | **Type of measure** | | **% Missing** | |
|---|---|---|---|---|
| | **Single item** | **Multiple item** | **Raw data** | **After missing value strategy** |
| Age | ✓ | | **11.5** | **11.5** |
| Gender | ✓ | | **0.7** | **0.7** |
| Ethnicity | ✓ | | **2.0** | **2.0** |
| Relationship status | ✓ | | **2.8** | **2.8** |
| Band | ✓ | | **0.9** | **0.9** |
| Years Nursing experience | ✓ | | **0.7** | **0.7** |
| No. contacted Hours | ✓ | | **8.5** | **8.5** |
| Full/Part time | ✓ | | **0.7** | **0.7** |
| Child Care responsibilities | ✓ | | **6.3** | **6.3** |
| Adult Care responsibilities | ✓ | | **6.9** | **6.9** |
| MBI Emotional Exhaustion | | ✓ | **2.6** | **0.4** |
| MBI Depersonalisation | | ✓ | **2.4** | **0.2** |
| MBI Personal Accomplishment | | ✓ | **3.3** | **0.4** |
| GHQ-12 | | ✓ | **3.7** | **0.0** |
| PCL-5 | | ✓ | **3.9** | **0.0** |
| Detachment after work | | ✓ | **0.4** | **0.0** |
| Recovery after work | | ✓ | **1.5** | **0.2** |
| Pace and amount of work | | ✓ | **1.3** | **0.2** |
| Emotional Load | | ✓ | **0.8** | **0.2** |
| Mental Load | | ✓ | **0.4** | **0.2** |
| Physical effort | | ✓ | **0.2** | **0.0** |
| Complexity of work | | ✓ | **0.2** | **0.0** |
| Role conflict | | ✓ | **1.3** | **0.4** |
| Work organisation | | ✓ | **1.7** | **0.2** |
| Disproportional expectations from relatives/visitors | | ✓ | **1.9** | **0.2** |
| Verbal aggression from relatives/visitors | | ✓ | **1.3** | **0.0** |
| Communication with Relatives | | ✓ | **0.9** | **0.2** |
| Task clarity | | ✓ | **0.6** | **0.2** |
| Autonomy | | ✓ | **1.3** | **0.2** |
| Feedback | | ✓ | **0.6** | **0.2** |
| Relationship with Supervisor | | ✓ | **1.5** | **0.2** |
| Relationship with Colleagues | | ✓ | **1.1** | **0.0** |
| Staffing | | ✓ | **0.2** | **0.2** |
| Focus on wellbeing | | ✓ | **1.3** | **0.2** |
| Effectiveness in goals | | ✓ | **0.4** | **0.2** |
| Quality | | ✓ | **0.2** | **0.2** |
| Opportunities to learn | | ✓ | **0.2** | **0.2** |
| Resilience | | ✓ | **1.1** | **0.2** |
| UWES Vigour | | ✓ | **1.3** | **0.7** |
| UWES Dedication | | ✓ | **1.1** | **0.2** |
| UWES Absorption | | ✓ | **1.3** | **0.2** |
| Changing Jobs | | ✓ | **0.6** | **0.2** |
| Certainty about future (at work) | | * | **0.0** | **0.0** |
| Patient safety | ✓ | | **5.2** | **5.2** |
| Quality of care | ✓ | | **6.1** | **6.1** |
| Satisfaction with organisation | ✓ | | **0.4** | **0.4** |
| Commitment to Organisation | | ✓ | **2.0** | **0.0** |

### REGRESSION ANALYSES

Categorical data were coded as follows:
Band: 0=band 5, 1=band 6+

Childcare responsibilities: 0=no, 1=yes

Adult care responsibilities: 0=no, 1=yes

Gender: 0=female, 1=male

Relationship status: 0=in steady relationship/married; 1=single/widowed/divorced separated

Full/Part time: Part time =0, Full time=1

#### PREDICTORS OF HEALTH IMPAIRMENT

#### Table A3: GHQ-12

**Dependent Variable: GHQ-12**

| | | Unstandardized Coefficients | | Standardized Coefficients | | | |
|---|---|---|---|---|---|---|---|
| Model | Predictors | B | Std Error | Beta | t | Sig· | $\Delta R^2$ |
| Step 1 | | | | | | | 2·8** |
| | Nurse experience (yrs) | -·10 | ·03 | -·16 | -3·13 | ·002 | |
| | Band | -·43 | ·72 | -·03 | -·60 | ·55 | |
| Step 2 | | | | | | | 23·5*** |
| | Nurse experience (yrs) | -·06 | ·03 | -·09 | -2·05 | ·04 | |
| | Band | -1·81 | ·66 | -·13 | -2·76 | ·01 | |
| | Pace and amount work | ·06 | ·02 | ·14 | 2·63 | ·01 | |
| | Emotional Load | ·11 | ·03 | ·25 | 4·44 | <·001 | |
| | Mental Load | ·00 | ·02 | ·01 | ·14 | ·89 | |
| | Physical effort | -·01 | ·02 | -·02 | -·36 | ·72 | |
| | Complexity of work | ·02 | ·02 | ·05 | ·93 | ·35 | |
| | Problems with Role | ·02 | ·03 | ·05 | ·93 | ·35 | |
| | Work Organisation | ·04 | ·03 | ·08 | 1·50 | ·14 | |
| | Relatives Expectations | ·72 | ·72 | ·06 | 1·00 | ·32 | |
| | Relatives Verbal Aggression | -·62 | ·71 | -·06 | -·87 | ·38 | |
| | Communication with Relatives | ·50 | ·16 | ·14 | 3·23 | ·001 | |

*p<0·05, **p<0·01 *** p<0·001

**Table A4: MBI: Emotional Exhaustion**

| Dependent Variable: MBI Emotional Exhaustion | | | | | | | |
|---|---|---|---|---|---|---|---|
| | | Unstandardized Coefficients | | Standardized Coefficients | | | · |
| Model | Predictors | B | Std Error | Beta | t | Sig· | $\Delta R^2$ |
| Step 1 | | | | | | | 6·5*** |
| | Nurse experience (yrs) | -0·18 | 0·06 | -0·16 | -3·30 | 0·001 | |
| | Childcare responsibilities | -3·44 | 1·16 | -0·15 | -2·97 | 0·003 | |
| | Relationship status | 1·95 | 1·43 | 0·07 | 1·36 | 0·17 | |
| Step 2 | | | | | | | 38·2*** |
| | Nurse experience (yrs) | -0·10 | 0·04 | -0·09 | -2·20 | 0·03 | |
| | Childcare responsibilities | -2·64 | 0·91 | -0·11 | -2·91 | 0·004 | |
| | Relationship status | 0·96 | 1·14 | 0·03 | 0·85 | 0·40 | |
| | Pace and amount work | 0·14 | 0·04 | 0·20 | 3·89 | <·001 | |
| | Emotional Load | 0·11 | 0·04 | 0·15 | 2·99 | 0·003 | |
| | Mental Load | 0·03 | 0·03 | 0·03 | 0·82 | 0·41 | |
| | Physical effort | 0·03 | 0·02 | 0·05 | 1·09 | 0·28 | |
| | Complexity of work | 0·05 | 0·03 | 0·08 | 1·77 | 0·08 | |
| | Problems with Role | 0·06 | 0·04 | 0·06 | 1·36 | 0·18 | |
| | Work Organisation | 0·16 | 0·04 | 0·18 | 3·99 | <·001 | |
| | Visitor Expectations | 3·45 | 1·08 | 0·18 | 3·19 | 0·002 | |
| | Visitor Verbal Aggression | -0·83 | 1·09 | -0·05 | -0·76 | 0·45 | |
| | Communication with Relatives | 0·29 | 0·23 | 0·05 | 1·25 | 0·21 | |

*p<0·05, **p<0·01 *** p<0·001

#### Table A5: MBI: Depersonalisation

**Dependent Variable: MBI Depersonalisation**

| Model | Predictors | Unstandardized Coefficients | | Standardized Coefficients | | | |
|---|---|---|---|---|---|---|---|
| | | B | Std· Error | Beta | t | Sig· | ΔR² |
| Step 1 | | | | | | | 6·6*** |
| | Nurse experience (yrs) | -·10 | ·03 | -·18 | -3·44 | <·001 | |
| | Nº contracted hours | ·02 | ·05 | ·02 | ·35 | ·72 | |
| | Childcare responsibilities | -1·15 | ·62 | -·10 | -1·87 | ·06 | |
| | Relationship status | 1·24 | ·74 | ·09 | 1·68 | ·09 | |
| Step 2 | | | | | | | 22·8*** |
| | Nurse experience (yrs) | -·08 | ·03 | -·14 | -2·98 | ·003 | |
| | Nº·contracted hours | ·00 | ·05 | ·00 | ·05 | ·96 | |
| | Childcare responsibilities | -·88 | ·55 | -·08 | -1·60 | ·11 | |
| | Relationship status | 1·10 | ·67 | ·08 | 1·66 | ·10 | |
| | Pace and amount work | ·01 | ·02 | ·03 | ·44 | ·66 | |
| | Emotional Load | ·06 | ·02 | ·15 | 2·59 | ·01 | |
| | Mental Load | -·02 | ·02 | -·05 | -·97 | ·33 | |
| | Physical effort | ·01 | ·01 | ·03 | ·59 | ·56 | |
| | Complexity of work | ·01 | ·02 | ·02 | ·32 | ·75 | |
| | Problems with Role | ·09 | ·02 | ·20 | 3·68 | <·001 | |
| | Work Organisation | ·05 | ·02 | ·12 | 2·26 | ·02 | |
| | Relatives Expectations | 1·58 | ·64 | ·17 | 2·49 | ·01 | |
| | Relatives Verbal Aggression | -·25 | ·64 | -·03 | -·39 | ·70 | |
| | Communication with Relatives | ·05 | ·14 | ·02 | ·40 | ·69 | |

*p<0·05, **p<0·01 *** p<0·001

#### Table A6: MBI: Personal Achievement

**Dependent Variable: MBI Personal Achievement**

| Model | Predictors | Unstandardized Coefficients | | Standardized Coefficients | | | |
|---|---|---|---|---|---|---|---|
| | | B | Std· Error | Beta | t | Sig· | ΔR$^2$ |
| Step 1 | | | | | | | 6·0*** |
| | Nurse experience (yrs) | ·07 | ·04 | ·10 | 1·97 | ·05 | |
| | Full/Part time | 2·91 | 1·65 | ·17 | 1·77 | ·08 | |
| | Adult care responsibilities | -3·04 | 1·05 | -·15 | -2·89 | ·004 | |
| | No· contracted hours | ·04 | ·13 | ·03 | ·31 | ·76 | |
| Step 2 | | | | | | | 6·5** |
| | Nurse experience (yrs) | ·07 | ·04 | ·10 | 1·95 | ·05 | |
| | Full/Part time | 2·39 | 1·65 | ·14 | 1·45 | ·15 | |
| | Adult care responsibilities | -2·43 | 1·04 | -·12 | -2·35 | ·02 | |
| | No· contracted hours | ·07 | ·13 | ·06 | ·57 | ·57 | |
| | Pace and amount work | -·02 | ·03 | -·04 | -·65 | ·52 | |
| | Emotional Load | ·04 | ·03 | ·08 | 1·27 | ·21 | |
| | Mental Load | ·06 | ·03 | ·11 | 1·97 | ·05 | |
| | Physical effort | -·03 | ·02 | -·10 | -1·72 | ·09 | |
| | Complexity of work | -·05 | ·03 | -·13 | -2·04 | ·04 | |
| | Problems with Role | -·08 | ·03 | -·14 | -2·23 | ·03 | |
| | Work Organisation | ·06 | ·03 | ·10 | 1·69 | ·09 | |
| | Relatives Expectations | -·74 | ·91 | -·06 | -·82 | ·41 | |
| | Relatives Verbal Aggression | ·31 | ·90 | ·03 | ·35 | ·73 | |
| | Communication with Relatives | -·10 | ·19 | -·03 | -·52 | ·60 | |

*p<0·05, **p<0·01 *** p<0·001

#### Table A7: UWES: Vigour

**Dependent Variable: UWES Vigour**

| Model | Predictor | Unstandardized Coefficients B | Unstandardized Coefficients Std· Error | Standardized Coefficients Beta | t | Sig· | ΔR² |
|---|---|---|---|---|---|---|---|
| Step 1 | | | | | | | 2·2** |
| | Nurse experience (yrs) | 0·01 | 0·01 | 0·08 | 1·67 | 0·10 | |
| | Childcare responsibilities | 0·29 | 0·12 | 0·12 | 2·36 | 0·02 | |
| Step 2 | | | | | | | 31·3*** |
| | Nurse experience (yrs) | 0·00 | 0·01 | 0·01 | 0·20 | 0·85 | |
| | Childcare responsibilities | 0·13 | 0·10 | 0·05 | 1·28 | 0·20 | |
| | Task Clarity | 0·00 | 0·00 | -0·03 | -0·60 | 0·55 | |
| | Autonomy | 0·00 | 0·00 | -0·03 | -0·51 | 0·61 | |
| | Feedback | 0·00 | 0·00 | -0·07 | -1·38 | 0·17 | |
| | Relationship with supervisor | 0·00 | 0·00 | -0·05 | -1·04 | 0·30 | |
| | Relationship with colleagues | 0·00 | 0·00 | -0·03 | -0·52 | 0·60 | |
| | Staffing | 0·00 | 0·00 | -0·06 | -1·18 | 0·24 | |
| | Wellbeing | -0·01 | 0·00 | -0·09 | -1·87 | 0·06 | |
| | Effectiveness achieving goals | 0·01 | 0·00 | 0·08 | 1·49 | 0·14 | |
| | Quality | -0·01 | 0·00 | -0·15 | -2·73 | 0·01 | |
| | Learning Opportunities | -0·01 | 0·00 | -0·12 | -2·48 | 0·01 | |
| | Resilience | 0·76 | 0·09 | 0·38 | 8·33 | <·001 | |

*p<0·05, **p<0·01 *** p<0·001

**Table A8: UWES: Dedication**

**Dependent Variable: UWES Dedication**

| Model | Predictor | Unstandardized Coefficients B | Std· Error | Standardized Coefficients Beta | t | Sig· | ΔR$^2$ |
|---|---|---|---|---|---|---|---|
| Step 1 | | | | | | | 3·5** |
| | Childcare responsibilities | 0·34 | 0·13 | 0·14 | 2·66 | 0·01 | |
| | Nurse experience (yrs) | 0·01 | 0·01 | 0·08 | 1·57 | 0·12 | |
| | No· contracted hours | 0·03 | 0·01 | 0·15 | 2·80 | 0·01 | |
| Step 2 | | | | | | | 31·6*** |
| | Childcare responsibilities | 0·14 | 0·11 | 0·06 | 1·31 | 0·19 | |
| | Nurse experience (yrs) | 0·00 | 0·01 | 0·04 | 0·76 | 0·45 | |
| | No· contracted hours | 0·02 | 0·01 | 0·08 | 1·60 | 0·11 | |
| | Task Clarity | 0·00 | 0·00 | -0·06 | -1·17 | 0·24 | |
| | Autonomy | 0·00 | 0·00 | 0·05 | 0·97 | 0·34 | |
| | Feedback | 0·00 | 0·00 | -0·02 | -0·39 | 0·70 | |
| | Relationship with supervisor | -0·01 | 0·00 | -0·07 | -1·23 | 0·22 | |
| | Relationship with colleagues | 0·00 | 0·01 | -0·03 | -0·57 | 0·57 | |
| | Staffing | 0·00 | 0·00 | -0·06 | -1·28 | 0·20 | |
| | Wellbeing | 0·00 | 0·00 | -0·04 | -0·91 | 0·37 | |
| | Effectiveness achieving goals | 0·00 | 0·01 | -0·01 | -0·17 | 0·87 | |
| | Quality | -0·01 | 0·00 | -0·11 | -1·95 | 0·05 | |
| | Learning Opportunities | -0·01 | 0·00 | -0·24 | -4·64 | <·001 | |
| | Resilience | 0·65 | 0·10 | 0·33 | 6·84 | <·001 | |

*p<0·05, **p<0·01 *** p<0·001

**Table A9: UWES: Absorption**

**Dependent Variable: UWES Absorption**

| Model | Predictor | Unstandardized Coefficients B | Unstandardized Coefficients Std· Error | Standardized Coefficients Beta | t | Sig· | ΔR² |
|---|---|---|---|---|---|---|---|
| Step 1 | | | | | | | 3·8** |
| | Band | 0·13 | 0·14 | 0·05 | 0·95 | 0·35 | |
| | Nurse experience (yrs) | 0·02 | 0·01 | 0·13 | 2·37 | 0·02 | |
| | Full/Part time | 0·04 | 0·27 | 0·02 | 0·17 | 0·87 | |
| | No· contracted hours | 0·03 | 0·02 | 0·13 | 1·34 | 0·18 | |
| Step 2 | | | | | | | 24·4*** |
| | Band | -0·02 | 0·12 | -0·01 | -0·18 | 0·86 | |
| | Nurse experience (yrs) | 0·01 | 0·01 | 0·08 | 1·59 | 0·11 | |
| | Full/Part time | -0·06 | 0·24 | -0·02 | -0·23 | 0·82 | |
| | No· contracted hours | 0·02 | 0·02 | 0·11 | 1·23 | 0·22 | |
| | Task Clarity | 0·00 | 0·00 | 0·00 | -0·08 | 0·94 | |
| | Autonomy | 0·00 | 0·00 | -0·02 | -0·43 | 0·67 | |
| | Feedback | 0·00 | 0·00 | -0·07 | -1·42 | 0·16 | |
| | Relationship with supervisor | 0·00 | 0·00 | 0·00 | 0·00 | 1·00 | |
| | Relationship with colleagues | 0·00 | 0·01 | 0·05 | 0·88 | 0·38 | |
| | Staffing | 0·00 | 0·00 | 0·00 | 0·08 | 0·94 | |
| | Wellbeing | 0·00 | 0·00 | 0·00 | 0·03 | 0·97 | |
| | Effectiveness achieving goals | 0·01 | 0·01 | 0·07 | 1·21 | 0·23 | |
| | Quality | -0·02 | 0·00 | -0·24 | -4·06 | <·001 | |
| | Learning Opportunities | -0·01 | 0·00 | -0·13 | -2·58 | 0·01 | |
| | Resilience | 0·72 | 0·10 | 0·37 | 7·44 | <·001 | |

*p<0·05, **p<0·01 *** p<0·001

**PREDICTORS OF ORGANISATIONAL OUTCOMES**

**Table A10: Commitment**

**Dependent Variable: Commitment**

| Model | Predictor | Unstandardized Coefficients B | Std· Error | Standardized Coefficients Beta | t | Sig· | ΔR² |
|---|---|---|---|---|---|---|---|
| Step 1 | | | | | | | 5·4*** |
| | Nurse experience (yrs) | ·00 | ·09 | ·00 | -·01 | ·99 | |
| | Childcare responsibilities | -3·34 | 1·79 | -·09 | -1·87 | ·06 | |
| | Band | -7·83 | 1·96 | -·21 | -4·00 | <·001 | |
| Step 2 | | | | | | | 37·4*** |
| | Nurse experience (yrs) | ·07 | ·08 | ·04 | ·86 | ·39 | |
| | Childcare responsibilities | -1·36 | 1·46 | -·04 | -·93 | ·35 | |
| | Band | -5·41 | 1·70 | -·14 | -3·17 | ·002 | |
| | Pace and amount work | -·06 | ·06 | -·05 | -·99 | ·32 | |
| | Emotional Load | -·08 | ·06 | -·07 | -1·28 | ·20 | |
| | Mental Load | -·02 | ·06 | -·01 | -·27 | ·79 | |
| | Physical effort | ·06 | ·04 | ·07 | 1·42 | ·16 | |
| | Complexity of work | ·00 | ·05 | ·00 | ·08 | ·94 | |
| | Problems with Role | ·05 | ·07 | ·04 | ·70 | ·48 | |
| | Work Organisation | ·13 | ·07 | ·10 | 1·93 | ·05 | |
| | Relatives Expectations | 2·19 | 1·83 | ·07 | 1·20 | ·23 | |
| | Relatives Verbal Aggression | -2·06 | 1·83 | -·07 | -1·12 | ·26 | |
| | Communication with Relatives | ·32 | ·39 | ·03 | ·81 | ·42 | |
| | Resilience | -2·24 | 1·37 | -·07 | -1·63 | ·10 | |
| | Task Clarity | -·04 | ·05 | -·04 | -·86 | ·39 | |
| | Autonomy | ·05 | ·05 | ·05 | ·96 | ·34 | |
| | Feedback | ·10 | ·04 | ·11 | 2·24 | ·03 | |
| | Relationship with supervisor | -·01 | ·05 | -·01 | -·15 | ·88 | |
| | Relationship with colleagues | -·05 | ·07 | -·04 | -·80 | ·42 | |
| | Staffing | -·07 | ·05 | -·07 | -1·44 | ·15 | |
| | Learning Opportunities | ·12 | ·04 | ·15 | 3·09 | ·002 | |
| | Wellbeing | ·27 | ·04 | ·30 | 6·33 | <·001 | |
| | Effectiveness achieving goals | ·08 | ·06 | ·07 | 1·34 | ·18 | |
| | Quality | ·22 | ·06 | ·20 | 3·70 | <·001 | |
| Step 3 | | | | | | | 5·1*** |
| | Nurse experience (yrs) | ·03 | ·08 | ·02 | ·42 | ·67 | |
| | Childcare responsibilities | -1·20 | 1·44 | -·03 | -·83 | ·40 | |
| | Band | -6·12 | 1·69 | -·16 | -3·63 | <·001 | |
| | Pace and amount work | -·03 | ·06 | -·03 | -·52 | ·61 | |
| | Emotional Load | -·02 | ·06 | -·02 | -·30 | ·77 | |

| | | | | | |
|---|---|---|---|---|---|
| Mental Load | ·00 | ·06 | ·00 | -·08 | ·94 |
| Physical effort | ·05 | ·04 | ·06 | 1·28 | ·20 |
| Complexity of work | ·00 | ·05 | ·00 | -·09 | ·93 |
| Problems with Role | ·02 | ·07 | ·01 | ·26 | ·79 |
| Work Organisation | ·15 | ·07 | ·11 | 2·16 | ·03 |
| Relatives Expectations | 2·05 | 1·81 | ·07 | 1·13 | ·26 |
| Relatives Verbal Aggression | -2·74 | 1·81 | -·10 | -1·52 | ·13 |
| Communication with Relatives | ·48 | ·39 | ·05 | 1·24 | ·21 |
| Resilience | -·08 | 1·59 | ·00 | -·05 | ·96 |
| Task Clarity | -·05 | ·05 | -·05 | -1·11 | ·27 |
| Autonomy | ·04 | ·05 | ·04 | ·95 | ·34 |
| Feedback | ·09 | ·04 | ·09 | 2·01 | ·05 |
| Relationship with supervisor | -·01 | ·05 | -·01 | -·23 | ·82 |
| Relationship with colleagues | -·03 | ·07 | -·02 | -·40 | ·69 |
| Staffing | -·07 | ·04 | -·08 | -1·65 | ·10 |
| Learning Opportunities | ·09 | ·04 | ·11 | 2·25 | ·02 |
| Wellbeing | ·29 | ·04 | ·31 | 6·67 | <·001 |
| Effectiveness achieving goals | ·08 | ·06 | ·07 | 1·31 | ·19 |
| Quality | ·19 | ·06 | ·17 | 3·21 | ·001 |
| UWES Vigour | -·96 | ·92 | -·06 | -1·04 | ·30 |
| UWES Dedication | -·14 | 1·04 | -·01 | -·13 | ·90 |
| UWES Absorption | -2·44 | ·87 | -·17 | -2·82 | ·01 |
| GHQ-12 | ·11 | ·17 | ·04 | ·62 | ·53 |
| MBI Emotional Exhaustion | ·16 | ·12 | ·10 | 1·39 | ·17 |
| MBI Depersonalisation | -·08 | ·16 | -·03 | -·50 | ·62 |
| MBI Personal Achievement | -·02 | ·14 | -·01 | -·17 | ·87 |
| PCL-5 | -·18 | ·07 | -·18 | -2·61 | ·01 |
| Recovery from work | -·05 | ·05 | -·06 | -·95 | ·34 |
| Detachment from work | -·01 | ·05 | -·01 | -·17 | ·86 |

*p<0·05, **p<0·01 *** p<0·001

**Table A11: Changing Jobs**
**Dependent variable: Changing Jobs**

| Model | | Unstandardized Coefficients | | Standardized Coefficients | | | |
|---|---|---|---|---|---|---|---|
| | | B | Std. Error | Beta | t | Sig. | $\Delta R^2$ |
| Step 1 | | | | | | | 4.4*** |
| | Band | -6.76 | 2.98 | -.12 | -2.27 | .02 | |
| | Nurse experience (yrs) | -.30 | .14 | -.11 | -2.11 | .04 | |
| | Full/Part time | -7.57 | 3.14 | -.12 | -2.41 | .02 | |
| Step 2 | | | | | | | 25.2*** |
| | Band | -5.33 | 2.83 | -.09 | -1.88 | .06 | |
| | Nurse experience (yrs) | -.24 | .13 | -.09 | -1.76 | .08 | |
| | Full/Part time | -6.73 | 2.91 | -.11 | -2.31 | .02 | |
| | Pace and amount work | -.13 | .10 | -.07 | -1.23 | .22 | |
| | Emotional Load | .10 | .10 | .06 | 1.00 | .32 | |
| | Mental Load | -.07 | .09 | -.04 | -.79 | .43 | |
| | Physical effort | .07 | .06 | .05 | 1.06 | .29 | |
| | Complexity of work | -.11 | .08 | -.07 | -1.28 | .20 | |
| | Problems with Role | -.01 | .12 | .00 | -.09 | .93 | |
| | Work Organisation | .10 | .11 | .05 | .90 | .37 | |
| | Relatives Expectations | 2.36 | 3.04 | .05 | .78 | .44 | |
| | Relatives Verbal Aggression | 2.09 | 3.00 | .05 | .70 | .49 | |
| | Communication with Relatives | .85 | .66 | .06 | 1.29 | .20 | |
| | Resilience | -5.13 | 2.31 | -.11 | -2.22 | .03 | |
| | Task Clarity | .01 | .08 | .00 | .07 | .95 | |
| | Autonomy | .04 | .08 | .02 | .47 | .64 | |
| | Feedback | .03 | .07 | .02 | .44 | .66 | |
| | Relationship with supervisor | .25 | .09 | .16 | 2.87 | .004 | |
| | Relationship with colleagues | .18 | .11 | .09 | 1.65 | .10 | |
| | Staffing | -.02 | .08 | -.01 | -.24 | .81 | |
| | Learning Opportunities | .25 | .06 | .20 | 3.85 | <.001 | |
| | Wellbeing | .16 | .07 | .11 | 2.17 | .03 | |
| | Effectiveness achieving goals | -.03 | .10 | -.02 | -.30 | .76 | |
| | Quality | .04 | .10 | .03 | .45 | .65 | |
| 3 | | | | | | | 7.3*** |
| | Band | -4.35 | 2.76 | -.07 | -1.58 | .12 | |
| | Nurse experience (yrs) | -.23 | .13 | -.09 | -1.74 | .08 | |
| | Full/Part time | -6.70 | 2.84 | -.11 | -2.36 | .02 | |
| | Pace and amount work | -.16 | .10 | -.09 | -1.58 | .11 | |
| | Emotional Load | .02 | .11 | .01 | .15 | .88 | |
| | Mental Load | -.07 | .09 | -.04 | -.79 | .43 | |

| | | | | | |
|---|---|---|---|---|---|
| Physical effort | .07 | .06 | .06 | 1.15 | .25 |
| Complexity of work | -.12 | .08 | -.07 | -1.41 | .16 |
| Problems with Role | -.04 | .12 | -.02 | -.30 | .77 |
| Work Organisation | .00 | .11 | .00 | -.02 | .99 |
| Relatives Expectations | 1.29 | 2.99 | .03 | .43 | .67 |
| Relatives Verbal Aggression | 2.37 | 2.92 | .05 | .81 | .42 |
| Communication with Relatives | .32 | .64 | .02 | .50 | .62 |
| Resilience | .89 | 2.62 | .02 | .34 | .73 |
| Task Clarity | .02 | .08 | .01 | .21 | .83 |
| Autonomy | .06 | .08 | .04 | .78 | .43 |
| Feedback | .02 | .07 | .01 | .26 | .80 |
| Relationship with supervisor | .23 | .08 | .15 | 2.75 | .01 |
| Relationship with colleagues | .11 | .11 | .06 | 1.05 | .29 |
| Staffing | -.06 | .07 | -.04 | -.78 | .43 |
| Learning Opportunities | .17 | .06 | .14 | 2.76 | .01 |
| Wellbeing | .09 | .07 | .07 | 1.32 | .19 |
| Effectiveness achieving goals | .01 | .10 | .01 | .12 | .90 |
| Quality | .06 | .09 | .03 | .60 | .55 |
| UWES Vigour | -.89 | 1.51 | -.04 | -.59 | .55 |
| UWES Dedication | -6.07 | 1.69 | -.27 | -3.59 | <.001 |
| UWES Absorption | 1.39 | 1.45 | .06 | .96 | .34 |
| GHQ-12 | .70 | .28 | .17 | 2.47 | .01 |
| MBI Emotional Exhaustion | .39 | .19 | .16 | 2.06 | .04 |
| MBI Depersonalisation | -.44 | .27 | -.09 | -1.66 | .10 |
| MBI Personal Achievement | .31 | .22 | .08 | 1.42 | .16 |
| PCL-5 | -.15 | .11 | -.09 | -1.29 | .20 |
| Recovery from work | .10 | .09 | .08 | 1.12 | .26 |
| Detachment from work | -.06 | .07 | -.05 | -.84 | .40 |

**Table A12: Certainty about the Future**

**Dependent Variable: Certainty about future**

| Model | Predictor | Unstandardized Coefficients | | Standardized Coefficients | t | Sig· | $\Delta R^2$ |
|---|---|---|---|---|---|---|---|
| | | B | Std· Error | Beta | | | |
| Step 1 | | | | | | | 1·7* |
| | Nurse experience (yrs) | -·02 | ·12 | -·01 | -·19 | ·85 | |
| | Childcare responsibilities | -6·32 | 2·46 | -·13 | -2·57 | ·01 | |
| Step 2 | | | | | | | 24·1*** |
| | Nurse experience (yrs) | ·04 | ·12 | ·02 | ·32 | ·75 | |
| | Childcare responsibilities | -3·49 | 2·23 | -·07 | -1·57 | ·12 | |
| | Pace and amount work | ·07 | ·10 | ·05 | ·72 | ·47 | |
| | Emotional Load | ·02 | ·10 | ·01 | ·20 | ·84 | |
| | Mental Load | -·13 | ·09 | -·08 | -1·53 | ·13 | |
| | Physical effort | ·04 | ·06 | ·04 | ·75 | ·46 | |
| | Complexity of work | ·05 | ·08 | ·04 | ·67 | ·50 | |
| | Problems with Role | ·02 | ·11 | ·01 | ·14 | ·89 | |
| | Work Organisation | ·15 | ·10 | ·08 | 1·41 | ·16 | |
| | Relatives Expectations | -1·66 | 2·79 | -·04 | -·59 | ·55 | |
| | Relatives Verbal Aggression | ·86 | 2·80 | ·02 | ·31 | ·76 | |
| | Communication with Relatives | ·68 | ·60 | ·05 | 1·14 | ·26 | |
| | Resilience | -5·73 | 2·07 | -·14 | -2·77 | ·01 | |
| | Task Clarity | ·07 | ·08 | ·05 | ·92 | ·36 | |
| | Autonomy | ·11 | ·07 | ·08 | 1·47 | ·14 | |
| | Feedback | ·01 | ·07 | ·01 | ·16 | ·87 | |
| | Relationship with supervisor | ·02 | ·08 | ·02 | ·29 | ·77 | |
| | Relationship with colleagues | ·07 | ·10 | ·04 | ·72 | ·47 | |
| | Staffing | ·02 | ·07 | ·02 | ·29 | ·77 | |
| | Learning Opportunities | ·24 | ·06 | ·22 | 4·06 | <·001 | |
| | Wellbeing | ·17 | ·07 | ·14 | 2·64 | ·01 | |
| | Effectiveness achieving goals | -·12 | ·09 | -·08 | -1·30 | ·20 | |
| | Quality | ·01 | ·09 | ·01 | ·12 | ·90 | |
| Step 3 | | | | | | | 7·0*** |
| | Nurse experience (yrs) | ·07 | ·12 | ·03 | ·61 | ·54 | |
| | Childcare responsibilities | -2·66 | 2·20 | -·05 | -1·21 | ·23 | |
| | Pace and amount work | ·04 | ·09 | ·03 | ·43 | ·67 | |
| | Emotional Load | -·06 | ·10 | -·04 | -·65 | ·52 | |
| | Mental Load | -·12 | ·09 | -·07 | -1·43 | ·15 | |
| | Physical effort | ·04 | ·06 | ·04 | ·75 | ·46 | |
| | Complexity of work | ·03 | ·08 | ·02 | ·43 | ·67 | |
| | Problems with Role | -·03 | ·11 | -·02 | -·26 | ·80 | |

| | | | | | |
|---|---|---|---|---|---|
| Work Organisation | ·09 | ·11 | ·05 | ·87 | ·39 |
| Relatives Expectations | -2·15 | 2·76 | -·05 | -·78 | ·44 |
| Relatives Verbal Aggression | ·34 | 2·75 | ·01 | ·12 | ·90 |
| Communication with Relatives | ·25 | ·59 | ·02 | ·42 | ·68 |
| Resilience | 1·23 | 2·40 | ·03 | ·51 | ·61 |
| Task Clarity | ·08 | ·07 | ·06 | 1·03 | ·30 |
| Autonomy | ·13 | ·07 | ·10 | 1·82 | ·07 |
| Feedback | ·00 | ·07 | ·00 | -·06 | ·95 |
| Relationship with supervisor | ·00 | ·08 | ·00 | ·06 | ·95 |
| Relationship with colleagues | ·04 | ·10 | ·02 | ·40 | ·69 |
| Staffing | ·00 | ·07 | ·00 | -·05 | ·96 |
| Learning Opportunities | ·18 | ·06 | ·16 | 2·97 | ·003 |
| Wellbeing | ·13 | ·07 | ·10 | 1·91 | ·06 |
| Effectiveness achieving goals | -·09 | ·09 | -·05 | -·95 | ·34 |
| Quality | ·00 | ·09 | ·00 | -·04 | ·97 |
| UWES Vigour | -·37 | 1·41 | -·02 | -·26 | ·79 |
| UWES Dedication | -3·85 | 1·59 | -·19 | -2·42 | ·02 |
| UWES Absorption | -·67 | 1·33 | -·03 | -·50 | ·62 |
| GHQ-12 | ·67 | ·26 | ·19 | 2·55 | ·01 |
| MBI Emotional Exhaustion | ·28 | ·18 | ·13 | 1·56 | ·12 |
| MBI Depersonalisation | -·28 | ·24 | -·07 | -1·14 | ·26 |
| MBI Personal Achievement | ·11 | ·21 | ·03 | ·51 | ·61 |
| PCL-5 | -·07 | ·11 | -·05 | -·61 | ·54 |
| Recovery from work | ·05 | ·08 | ·05 | ·66 | ·51 |
| Detachment from work | -·01 | ·07 | -·01 | -·20 | ·84 |

*p<0·05, **p<0·01 *** p<0·001

**Table A13: Satisfaction with Organisation**

**Dependent Variable: Satisfaction with Organisation**

| | | Unstandardized Coefficients | | Standardized Coefficients | | | |
|---|---|---|---|---|---|---|---|
| Model | Predictor | B | Std· Error | Beta | t | Sig· | $\Delta R^2$ |
| Step 1 | | | | | | | 1·7* |
| | Nurse experience (yrs) | -·10 | ·12 | -·04 | -·81 | ·42 | |
| | Band | -5·52 | 2·60 | -·11 | -2·12 | ·03 | |
| Step 2 | | | | | | | 49·1*** |
| | Nurse experience (yrs) | ·04 | ·09 | ·02 | ·48 | ·63 | |
| | Band | -5·00 | 2·04 | -·10 | -2·45 | ·01 | |
| | Pace and amount work | ·09 | ·07 | ·06 | 1·17 | ·24 | |
| | Emotional Load | -·05 | ·08 | -·03 | -·70 | ·48 | |
| | Mental Load | ·05 | ·07 | ·03 | ·70 | ·49 | |
| | Physical effort | ·08 | ·05 | ·07 | 1·72 | ·09 | |
| | Complexity of work | ·03 | ·06 | ·02 | ·52 | ·61 | |
| | Problems with Role | -·07 | ·09 | -·04 | -·81 | ·42 | |
| | Work Organisation | ·08 | ·08 | ·05 | 1·01 | ·31 | |
| | Relatives Expectations | ·39 | 2·20 | ·01 | ·18 | ·86 | |
| | Relatives Verbal Aggression | ·47 | 2·18 | ·01 | ·21 | ·83 | |
| | Communication with Relatives | ·87 | ·48 | ·07 | 1·82 | ·07 | |
| | Resilience | -2·26 | 1·66 | -·06 | -1·36 | ·18 | |
| | Task Clarity | ·00 | ·06 | ·00 | ·01 | ·99 | |
| | Autonomy | -·02 | ·06 | -·01 | -·30 | ·77 | |
| | Feedback | ·09 | ·05 | ·07 | 1·70 | ·09 | |
| | Relationship with supervisor | ·09 | ·06 | ·07 | 1·46 | ·15 | |
| | Relationship with colleagues | ·08 | ·08 | ·05 | 1·04 | ·30 | |
| | Staffing | ·06 | ·05 | ·05 | 1·09 | ·28 | |
| | Learning Opportunities | ·14 | ·05 | ·13 | 2·96 | ·003 | |
| | Wellbeing | ·46 | ·05 | ·38 | 8·97 | <·001 | |
| | Effectiveness achieving goals | ·12 | ·07 | ·08 | 1·64 | ·10 | |
| | Quality | ·10 | ·07 | ·07 | 1·42 | ·16 | |
| Step 3 | | | | | | | 5·1*** |
| | Nurse experience (yrs) | ·11 | ·09 | ·04 | 1·12 | ·26 | |
| | Band | -5·26 | 1·99 | -·10 | -2·64 | ·01 | |
| | Pace and amount work | ·03 | ·07 | ·02 | ·41 | ·68 | |
| | Emotional Load | -·11 | ·08 | -·07 | -1·39 | ·16 | |
| | Mental Load | ·05 | ·07 | ·03 | ·79 | ·43 | |
| | Physical effort | ·09 | ·05 | ·08 | 1·87 | ·06 | |
| | Complexity of work | ·00 | ·06 | ·00 | -·01 | 1·00 | |
| | Problems with Role | -·08 | ·09 | -·05 | -·97 | ·33 | |

| | | | | | |
|---|---|---|---|---|---|
| Work Organisation | ·02 | ·08 | ·01 | ·18 | ·85 |
| Relatives Expectations | -·36 | 2·17 | -·01 | -·17 | ·87 |
| Relatives Verbal Aggression | ·05 | 2·12 | ·00 | ·02 | ·98 |
| Communication with Relatives | ·74 | ·47 | ·06 | 1·57 | ·12 |
| Resilience | 2·02 | 1·89 | ·05 | 1·07 | ·29 |
| Task Clarity | ·00 | ·06 | ·00 | ·00 | 1·00 |
| Autonomy | ·00 | ·06 | ·00 | ·04 | ·97 |
| Feedback | ·08 | ·05 | ·06 | 1·47 | ·14 |
| Relationship with supervisor | ·07 | ·06 | ·05 | 1·15 | ·25 |
| Relationship with colleagues | ·07 | ·08 | ·04 | ·83 | ·41 |
| Staffing | ·03 | ·05 | ·02 | ·51 | ·61 |
| Learning Opportunities | ·09 | ·05 | ·08 | 1·89 | ·06 |
| Wellbeing | ·42 | ·05 | ·34 | 8·21 | <·001 |
| Effectiveness achieving goals | ·15 | ·07 | ·09 | 2·05 | ·04 |
| Quality | ·09 | ·07 | ·07 | 1·37 | ·17 |
| UWES Vigour | -·24 | 1·09 | -·01 | -·22 | ·82 |
| UWES Dedication | -2·67 | 1·23 | -·14 | -2·18 | ·03 |
| UWES Absorption | -1·06 | 1·04 | -·05 | -1·01 | ·31 |
| GHQ-12 | -·19 | ·21 | -·05 | -·92 | ·36 |
| MBI Emotional Exhaustion | ·47 | ·14 | ·22 | 3·47 | <·001 |
| MBI Depersonalisation | -·11 | ·19 | -·03 | -·55 | ·58 |
| MBI Personal Achievement | ·07 | ·16 | ·02 | ·46 | ·64 |
| PCL-5 | -·06 | ·08 | -·05 | -·78 | ·44 |
| Recovery from work | ·02 | ·06 | ·02 | ·33 | ·74 |
| Detachment from work | ·10 | ·05 | ·09 | 1·78 | ·08 |

*p<0·05, **p<0·01 *** p<0·001

**Table A14: Patient Safety**

**Dependent Variable: Patient safety**

| | | Unstandardized Coefficients | | Standardized Coefficients | | | |
|---|---|---|---|---|---|---|---|
| Model | Predictor | B | Std· Error | Beta | t | Sig· | ΔR² |
| Step 1 | | | | | | | 1·5* |
| | Nurse experience (yrs) | ·01 | ·00 | ·12 | 2·53 | ·01 | |
| Step 2 | | | | | | | 21·5*** |
| | Nurse experience (yrs) | ·01 | ·00 | ·13 | 2·63 | ·01 | |
| | Pace and amount work | -·01 | ·00 | -·08 | -1·31 | ·19 | |
| | Emotional Load | ·01 | ·00 | ·11 | 1·79 | ·07 | |
| | Mental Load | ·00 | ·00 | ·01 | ·22 | ·83 | |
| | Physical effort | -·01 | ·00 | -·11 | -2·03 | ·04 | |
| | Complexity of work | ·00 | ·00 | -·03 | -·45 | ·65 | |
| | Problems with Role | ·00 | ·00 | ·01 | ·09 | ·93 | |
| | Work Organisation | ·00 | ·00 | ·03 | ·45 | ·65 | |
| | Relatives Expectations | ·26 | ·12 | ·16 | 2·23 | ·03 | |
| | Relatives Verbal Aggression | -·21 | ·12 | -·13 | -1·78 | ·08 | |
| | Communication with Relatives | -·05 | ·03 | -·10 | -2·06 | ·04 | |
| | Resilience | ·12 | ·09 | ·07 | 1·41 | ·16 | |
| | Task Clarity | ·00 | ·00 | -·03 | -·50 | ·62 | |
| | Autonomy | ·00 | ·00 | -·03 | -·58 | ·56 | |
| | Feedback | ·00 | ·00 | ·02 | ·39 | ·69 | |
| | Relationship with supervisor | ·00 | ·00 | -·06 | -1·01 | ·31 | |
| | Relationship with colleagues | ·00 | ·00 | -·05 | -·81 | ·42 | |
| | Staffing | ·00 | ·00 | -·06 | -1·02 | ·31 | |
| | Learning Opportunities | -·01 | ·00 | -·18 | -3·28 | ·001 | |
| | Wellbeing | ·00 | ·00 | ·00 | ·02 | ·98 | |
| | Effectiveness achieving goals | ·00 | ·00 | -·04 | -·70 | ·49 | |
| | Quality | -·01 | ·00 | -·13 | -2·13 | ·03 | |
| Step 3 | | | | | | | 3·6* |
| | Nurse experience (yrs) | ·01 | ·01 | ·09 | 1·72 | ·09 | |
| | Pace and amount work | ·00 | ·00 | -·06 | -·91 | ·36 | |
| | Emotional Load | ·01 | ·00 | ·12 | 1·90 | ·06 | |
| | Mental Load | ·00 | ·00 | ·01 | ·17 | ·86 | |
| | Physical effort | -·01 | ·00 | -·11 | -2·04 | ·04 | |
| | Complexity of work | ·00 | ·00 | -·04 | -·64 | ·52 | |
| | Problems with Role | ·00 | ·00 | ·02 | ·26 | ·79 | |
| | Work Organisation | ·00 | ·00 | ·06 | 1·04 | ·30 | |
| | Relatives Expectations | ·31 | ·12 | ·19 | 2·59 | ·01 | |
| | Relatives Verbal Aggression | -·20 | ·12 | -·12 | -1·67 | ·09 | |

| | | | | | |
|---|---|---|---|---|---|
| Communication with Relatives | -·06 | ·03 | -·12 | -2·49 | ·01 |
| Resilience | ·15 | ·10 | ·09 | 1·46 | ·14 |
| Task Clarity | ·00 | ·00 | -·03 | -·61 | ·54 |
| Autonomy | ·00 | ·00 | -·03 | -·62 | ·54 |
| Feedback | ·00 | ·00 | ·02 | ·40 | ·69 |
| Relationship with supervisor | ·00 | ·00 | -·05 | -·79 | ·43 |
| Relationship with colleagues | ·00 | ·00 | -·05 | -·90 | ·37 |
| Staffing | ·00 | ·00 | -·04 | -·78 | ·44 |
| Learning Opportunities | -·01 | ·00 | -·19 | -3·41 | <·001 |
| Wellbeing | ·00 | ·00 | ·00 | ·05 | ·96 |
| Effectiveness achieving goals | ·00 | ·00 | -·03 | -·58 | ·56 |
| Quality | -·01 | ·00 | -·13 | -2·03 | ·04 |
| UWES Vigour | ·09 | ·06 | ·11 | 1·52 | ·13 |
| UWES Dedication | -·05 | ·07 | -·07 | -·81 | ·42 |
| UWES Absorption | ·03 | ·06 | ·04 | ·54 | ·59 |
| GHQ-12 | ·03 | ·01 | ·21 | 2·70 | ·01 |
| MBI Emotional Exhaustion | ·00 | ·01 | -·03 | -·39 | ·69 |
| MBI Depersonalisation | -·03 | ·01 | -·15 | -2·47 | ·01 |
| MBI Personal Achievement | -·01 | ·01 | -·08 | -1·24 | ·22 |
| PCL-5 | ·00 | ·00 | -·08 | -·97 | ·33 |
| Recovery from work | ·00 | ·00 | ·04 | ·57 | ·57 |
| Detachment from work | ·00 | ·00 | -·10 | -1·52 | ·13 |

*p<0·05, **p<0·01 *** p<0·001

**Table A15: Quality of Care**

| **Dependent Variable: Quality of Care** | | | | | | | |
|---|---|---|---|---|---|---|---|
| | | Unstandardized Coefficients | | Standardized Coefficients | | | |
| Model | Predictor | B | Std· Error | Beta | t | Sig· | $\Delta R^2$ |
| 1 | | | | | | | 3·6*** |
| | Nurse experience (yrs) | ·01 | ·00 | ·10 | 1·97 | ·05 | |
| | Relationship status | -·37 | ·12 | -·15 | -3·07 | ·002 | |
| 2 | | | | | | | 24·4*** |
| | Nurse experience (yrs) | ·01 | ·00 | ·11 | 2·20 | ·03 | |
| | Relationship status | -·30 | ·11 | -·12 | -2·69 | ·01 | |
| | Pace and amount work | ·00 | ·00 | -·03 | -·54 | ·59 | |
| | Emotional Load | ·01 | ·00 | ·13 | 2·23 | ·03 | |
| | Mental Load | ·00 | ·00 | -·04 | -·72 | ·47 | |
| | Physical effort | ·00 | ·00 | -·04 | -·71 | ·48 | |
| | Complexity of work | ·00 | ·00 | -·05 | -·82 | ·41 | |
| | Problems with Role | ·00 | ·00 | ·03 | ·48 | ·63 | |
| | Work Organisation | -·01 | ·00 | -·08 | -1·37 | ·17 | |
| | Relatives Expectations | ·23 | ·11 | ·15 | 2·10 | ·04 | |
| | Relatives Verbal Aggression | -·33 | ·11 | -·22 | -3·02 | ·003 | |
| | Communication with Relatives | -·02 | ·02 | -·05 | -1·02 | ·31 | |
| | Resilience | -·03 | ·08 | -·02 | -·33 | ·75 | |
| | Task Clarity | ·00 | ·00 | -·04 | -·69 | ·49 | |
| | Autonomy | ·00 | ·00 | ·01 | ·25 | ·80 | |
| | Feedback | ·00 | ·00 | ·03 | ·54 | ·59 | |
| | Relationship with supervisor | ·00 | ·00 | -·02 | -·27 | ·78 | |
| | Relationship with colleagues | -·01 | ·00 | -·09 | -1·44 | ·15 | |
| | Staffing | -·01 | ·00 | -·12 | -2·31 | ·02 | |
| | Learning Opportunities | -·01 | ·00 | -·17 | -3·25 | ·001 | |
| | Wellbeing | ·00 | ·00 | ·08 | 1·45 | ·15 | |
| | Effectiveness achieving goals | ·00 | ·00 | -·03 | -·54 | ·59 | |
| | Quality | -·01 | ·00 | -·19 | -3·18 | ·002 | |
| 3 | | | | | | | 3·1 ns |
| | Nurse experience (yrs) | ·01 | ·00 | ·09 | 1·83 | ·07 | |
| | Relationship status | -·28 | ·11 | -·12 | -2·52 | ·01 | |
| | Pace and amount work | ·00 | ·00 | -·04 | -·58 | ·56 | |
| | Emotional Load | ·01 | ·00 | ·11 | 1·82 | ·07 | |
| | Mental Load | ·00 | ·00 | -·05 | -1·02 | ·31 | |
| | Physical effort | ·00 | ·00 | -·03 | -·66 | ·51 | |
| | Complexity of work | ·00 | ·00 | -·05 | -·95 | ·34 | |
| | Problems with Role | ·00 | ·00 | ·05 | ·83 | ·41 | |

| | | | | | |
|---|---|---|---|---|---|
| Work Organisation | ·00 | ·00 | -·06 | -·92 | ·36 |
| Relatives Expectations | ·26 | ·11 | ·17 | 2·36 | ·02 |
| Relatives Verbal Aggression | -·31 | ·11 | -·21 | -2·82 | ·01 |
| Communication with Relatives | -·03 | ·02 | -·07 | -1·40 | ·16 |
| Resilience | ·00 | ·10 | ·00 | -·05 | ·96 |
| Task Clarity | ·00 | ·00 | -·03 | -·58 | ·56 |
| Autonomy | ·00 | ·00 | ·01 | ·22 | ·83 |
| Feedback | ·00 | ·00 | ·04 | ·67 | ·50 |
| Relationship with supervisor | ·00 | ·00 | -·01 | -·23 | ·82 |
| Relationship with colleagues | -·01 | ·00 | -·09 | -1·50 | ·13 |
| Staffing | -·01 | ·00 | -·12 | -2·25 | ·03 |
| Learning Opportunities | -·01 | ·00 | -·17 | -3·04 | ·003 |
| Wellbeing | ·00 | ·00 | ·07 | 1·29 | ·20 |
| Effectiveness achieving goals | ·00 | ·00 | -·03 | -·48 | ·63 |
| Quality | -·01 | ·00 | -·17 | -2·83 | ·005 |
| UWES Vigour | ·06 | ·06 | ·08 | 1·08 | ·28 |
| UWES Dedication | ·05 | ·06 | ·07 | ·84 | ·40 |
| UWES Absorption | -·01 | ·05 | -·01 | -·10 | ·92 |
| GHQ-12 | ·02 | ·01 | ·15 | 1·97 | ·05 |
| MBI Emotional Exhaustion | ·00 | ·01 | ·05 | ·61 | ·54 |
| MBI Depersonalisation | -·02 | ·01 | -·14 | -2·26 | ·02 |
| MBI Personal Achievement | -·01 | ·01 | -·08 | -1·21 | ·23 |
| PCL-5 | ·00 | ·00 | ·02 | ·29 | ·77 |
| Recovery from work | ·00 | ·00 | -·06 | -·80 | ·42 |
| Detachment from work | ·00 | ·00 | ·01 | ·11 | ·92 |

*p<0·05, **p<0·01 *** p<0·001

#### MODERATION ANALYSES

Simple Slope Plots

**Figure A1:** Relationship between Job-Demands and Mental Health (GHQ-12): moderation by Relationship with Colleagues

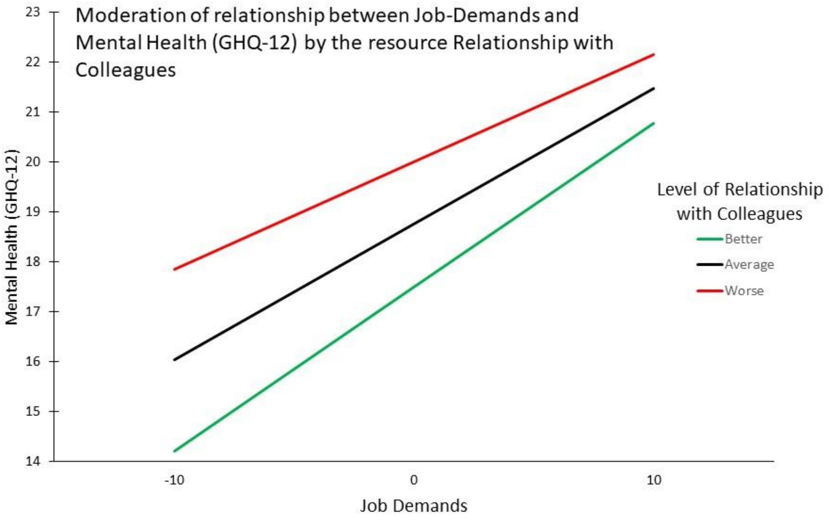

**Figure A2:** Relationship between Job-Demands and Burnout (MBI, Emotional Exhaustion): moderation by Learning Opportunities

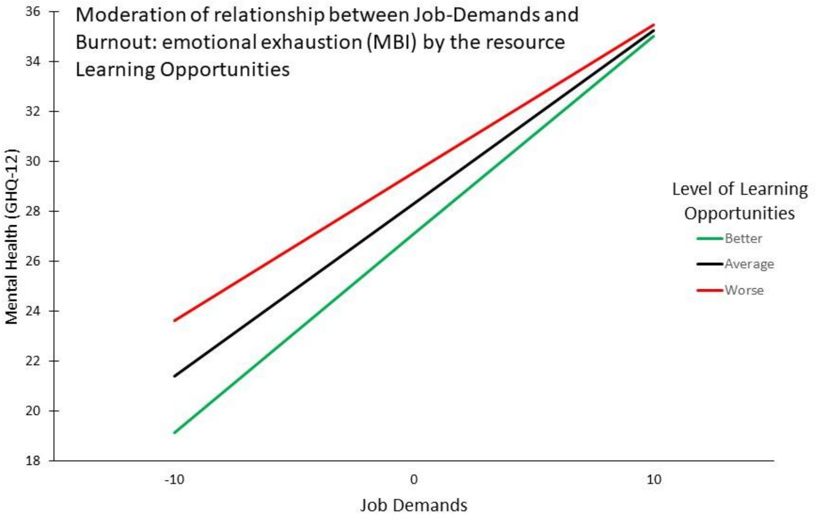

**Figure A3:** Relationship between Job-Demands and Burnout (MBI, Emotional Exhaustion): moderation by Prioritisation of Wellbeing by Employing Organisation

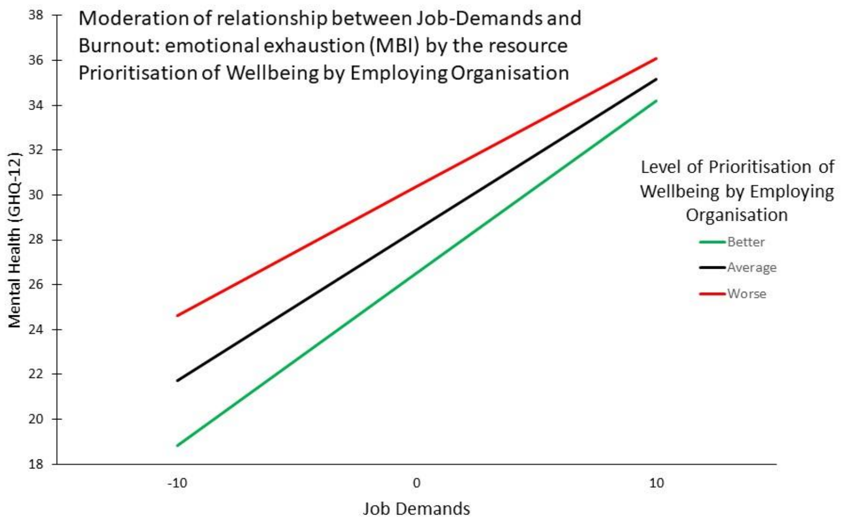

**Figure A4:** Relationship between Job-Demands and Burnout (MBI, Emotional Exhaustion): moderation by Relationship with Colleagues

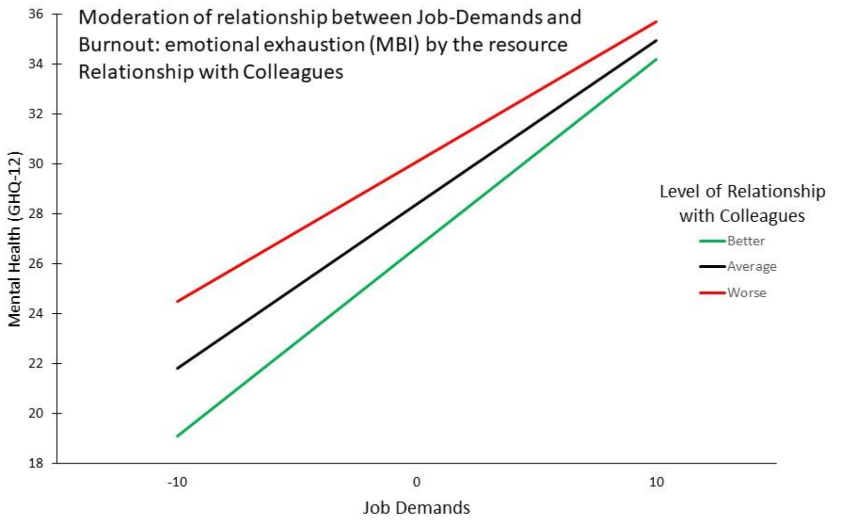

**Figure A5:** Relationship between Job-Demands and Burnout (MBI, Emotional Exhaustion): moderation by Relationship with Supervisor

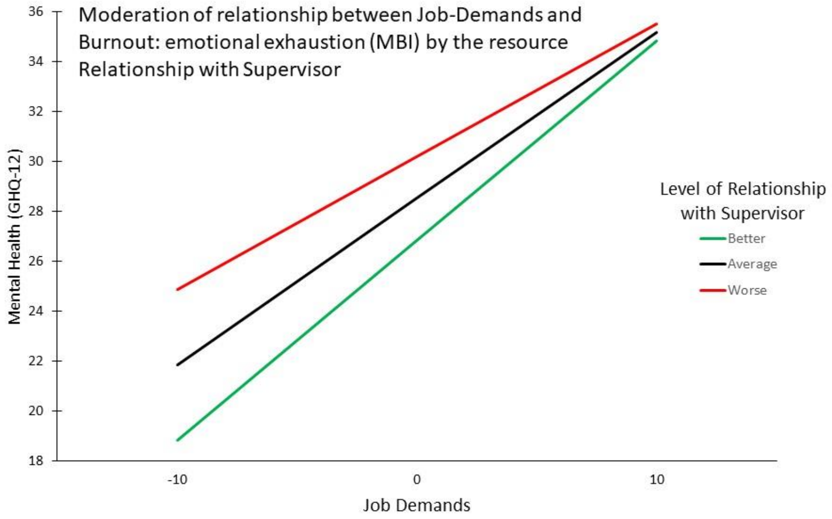

**Figure A6:** Relationship between Job-Demands and PTSD Symptoms (PCL-5): moderation by Learning Opportunities

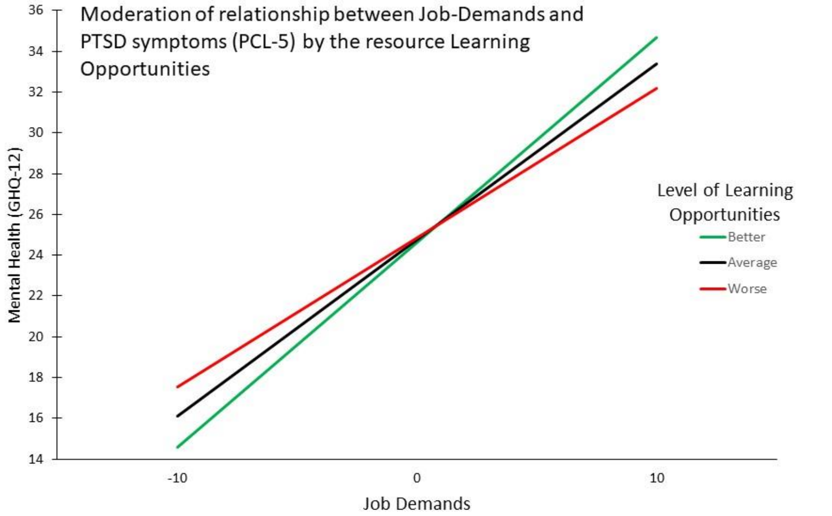
